# Vitamin D supplementation to prevent acute respiratory infections: systematic review and meta-analysis of aggregate data from randomised controlled trials

**DOI:** 10.1101/2020.07.14.20152728

**Authors:** David A Jolliffe, Carlos A Camargo, John D Sluyter, Mary Aglipay, John F Aloia, Davaasambuu Ganmaa, Peter Bergman, Arturo Borzutzky, Camilla T Damsgaard, Gal Dubnov-Raz, Susanna Esposito, Clare Gilham, Adit A Ginde, Inbal Golan-Tripto, Emma C Goodall, Cameron C Grant, Christopher J Griffiths, Anna Maria Hibbs, Wim Janssens, Anuradha Vaman Khadilkar, Ilkka Laaksi, Margaret T Lee, Mark Loeb, Jonathon L Maguire, Paweł Majak, David T Mauger, Semira Manaseki-Holland, David R Murdoch, Akio Nakashima, Rachel E Neale, Hai Pham, Christine Rake, Judy R Rees, Jenni Rosendahl, Robert Scragg, Dheeraj Shah, Yoshiki Shimizu, Steve Simpson-Yap, Geeta Trilok Kumar, Mitsuyoshi Urashima, Adrian R Martineau

## Abstract

**Background:** A 2017 meta-analysis of data from 25 randomised controlled trials of vitamin D supplementation for the prevention of acute respiratory infections revealed a protective effect of the intervention. Since then, 20 new RCTs have been completed.

**Methods:** Systematic review and meta-analysis of data from randomised controlled trials (RCTs) of vitamin D for ARI prevention using a random effects model. Pre-specified sub-group analyses were done to determine whether effects of vitamin D on risk of ARI varied according to baseline 25-hydroxyvitamin D (25[OH]D) concentration or dosing regimen. We searched MEDLINE, EMBASE, the Cochrane Central Register of Controlled Trials (CENTRAL), Web of Science and the ClinicalTrials.gov registry from inception to 1st May 2020. Double-blind RCTs of supplementation with vitamin D or calcidiol, of any duration, were eligible if they were approved by a Research Ethics Committee and if ARI incidence was collected prospectively and pre-specified as an efficacy outcome. Aggregate data, stratified by baseline 25(OH)D concentration, were obtained from study authors. The study was registered with PROSPERO (no. CRD42020190633).

**Findings:** We identified 45 eligible RCTs (total 73,384 participants). Data were obtained for 46,331 (98.0%) of 47,262 participants in 42 studies, aged 0 to 95 years. For the primary comparison of vitamin D supplementation vs. placebo, the intervention reduced risk of ARI overall (Odds Ratio [OR] 0.91, 95% CI 0.84 to 0.99; P for heterogeneity 0.01). No statistically significant effect of vitamin D was seen for any of the sub-groups defined by baseline 25(OH)D concentration. However, protective effects were seen for trials in which vitamin D was given using a daily dosing regimen (OR 0.75, 95% CI 0.61 to 0.93); at daily dose equivalents of 400-1000 IU (OR 0.70, 95% CI 0.55 to 0.89); and for a duration of ≤12 months (OR 0.82, 95% CI 0.72 to 0.93). No significant interaction was seen between allocation to vitamin D vs. placebo and dose frequency, dose size, or study duration. Vitamin D did not influence the proportion of participants experiencing at least one serious adverse event (OR 0.97, 95% CI 0.86 to 1.09). Risk of bias within individual studies was assessed as being low for all but three trials. A funnel plot showed left-sided asymmetry (P=0.008, Egger’s test).

**Interpretation:** Vitamin D supplementation was safe and reduced risk of ARI, despite evidence of significant heterogeneity across trials. Protection was associated with administration of daily doses of 400-1000 IU vitamin D for up to 12 months. The relevance of these findings to COVID-19 is not known and requires investigation.

**Funding:** None

## Research in context

### Evidence before this study

The active vitamin D metabolite, 1,25-dihydroxyvitamin D, induces innate immune responses to respiratory viruses and bacteria. A previous meta-analysis of individual participant data from 10,933 participants in 25 randomised controlled trials of vitamin D supplementation for the prevention of acute respiratory infection demonstrated an overall protective effect (adjusted Odds Ratio [aOR] 0.88, 95% confidence interval 0.81 to 0.96). Sub-group analysis revealed most benefit in those with the lowest vitamin D status at baseline who received daily or weekly supplementation (aOR 0.30, 0.17 to 0.53).

### Added value of this study

Our meta-analysis of aggregate data from 46,331 participants in 42 randomised controlled trials, stratified by baseline 25(OH)D concentration, provides an updated estimate of the protective effects of vitamin D against acute respiratory infection overall, and in sub-groups defined by baseline vitamin D status and dosing frequency, amount and duration.

### Implications of all the available evidence

Overall, vitamin D reduced the risk of having one or more acute respiratory infections (OR 0.91, 0.84 to 0.99), but there was evidence of significant heterogeneity across trials (P for heterogeneity 0.01). A funnel plot showed left-sided asymmetry, which may reflect publication bias and/or heterogeneity of effect across trials. No statistically significant effect of vitamin D was seen for any of the sub-groups defined by baseline 25(OH)D concentration. However, protective effects were seen in trials where vitamin D was given using a daily dosing regimen (OR 0.75, 0.61 to 0.93); at daily dose equivalents of 400-1000 IU (OR 0.70, 0.55 to 0.89); and for a duration of ≤12 months (OR 0.82, 0.72 to 0.93). The relevance of these findings to COVID-19 is not known and requires investigation.

## Introduction

Interest in the potential for vitamin D supplementation to reduce risk of acute respiratory infections (ARI) has increased since the emergence of the COVID-19 pandemic.^1^ This stems from findings of laboratory studies, showing that vitamin D metabolites support innate immune responses to respiratory viruses,^2^ together with observational studies reporting independent associations between low circulating levels of 25-hydroxyvitamin D (25[OH]D, the widely accepted biomarker of vitamin D status) and increased risk of ARI caused by other pathogens.^3,4^ Randomised controlled trials (RCTs) of vitamin D for the prevention of ARI have produced heterogeneous results, with some showing protection, and others reporting null findings. We previously meta-analysed individual participant data from 10,933 participants in 25 RCTs^5-29^ and showed a protective overall effect that was stronger in those with lower baseline 25(OH)D levels, and in trials where vitamin D was administered daily or weekly rather than in more widely spaced bolus doses.^30^ Since the date of the final literature search performed for that study (December 2015), 20 RCTs with 62,063 participants fulfilling the same eligibility criteria have been completed.^31-50^ We therefore sought data from these more recent studies for inclusion in an updated meta-analysis of stratified aggregate (trial-level) data to determine whether vitamin D reduced ARI risk overall, and to evaluate whether effects of vitamin D on ARI risk varied according to baseline 25(OH)D concentration and/or dosing regimen (frequency, dose size, and trial duration).

## Methods

### Protocol, Registration and Ethical Approvals

Methods were pre-specified in a protocol that was registered with the PROSPERO International Prospective Register of Systematic Reviews (https://www.crd.york.ac.uk/PROSPERO/display_record.php?RecordID=190633). Research Ethics Committee approval to conduct this meta-analysis was not required in the UK; local ethical permission to contribute data from primary trials was required and obtained for studies by Camargo *et al*^13^ (The Ethics Review Committee of the Mongolian Ministry of Health), Murdoch *et al*^14^ (Southern Health and Disability Ethics Committee, ref. URB/09/10/050/AM02), Rees *et al*^17^ (Committee for the Protection of Human Subjects, Dartmouth College, USA; Protocol # 24381), Tachimoto *et al*^28^ (Ethics committee of the Jikei University School of Medicine, ref 26-333: 7839), Tran *et al*^18^ (QIMR Berghofer Medical Research Institute Human Research Ethics Committee, P1570) and Urashima *et al*^6,20^ (Ethics committee of the Jikei University School of Medicine, ref 26-333: 7839).

### Eligibility Criteria

Randomised, double-blind, trials of supplementation with vitamin D_3_, vitamin D_2_ or 25(OH)D of any duration, with a placebo or low-dose vitamin D control, were eligible for inclusion if they had been approved by a Research Ethics Committee and if data on incidence of ARI were collected prospectively and pre-specified as an efficacy outcome. The latter requirement was imposed in order to minimise misclassification bias (prospectively designed instruments to capture ARI events were deemed more likely to be sensitive and specific for this outcome). Studies reporting results of long-term follow-up of primary RCTs were excluded.

### Study Identification and Selection

Two investigators (ARM and DAJ) searched MEDLINE, EMBASE, the Cochrane Central Register of Controlled Trials (CENTRAL), Web of Science and the ClinicalTrials.gov registry using the electronic search strategies described in the Methods Section of Supplementary Material. Searches were regularly updated up to and including 1^st^ May 2020. No language restrictions were imposed. These searches were supplemented by searching review articles and reference lists of trial publications. Collaborators were asked if they knew of any additional trials. Three investigators (DAJ, CAC and ARM) determined which trials met the eligibility criteria.

### Data Collection Processes

Summary data from trials which contributed to our previous meta-analysis of individual participant data^30^ were extracted from our central database, with permission from the Principal Investigators. Summary data relating to the primary outcome (overall and by sub-group) and secondary outcomes (overall only) from newly identified trials were requested from Principal Investigators. On receipt, they were assessed for consistency with associated publications. Study authors were contacted to provide missing data and to resolve any queries arising from these consistency checks. Once queries had been resolved, clean summary data were uploaded to the study database, which was held in STATA IC v14.2 (StataCorp, College Station, TX).

Data relating to study characteristics were extracted for the following variables: study setting, eligibility criteria, 25(OH)D assay and levels, details of intervention and control regimens, trial duration, case definitions for ARI and number entering primary analysis (after randomisation). Follow-up summary data were requested for the proportions of participants experiencing one or more ARI during the trial, both overall and stratified by baseline serum 25(OH)D concentration, where this was available. We also requested summary data on the proportions of participants who experienced one or more of the following events during the trial: upper respiratory infection (URI); lower respiratory infection (LRI); Emergency Department attendance and/or hospital admission for ARI; death due to ARI or respiratory failure; use of antibiotics to treat an ARI; absence from work or school due to ARI; a serious adverse event; death due to any cause; and potential adverse reactions to vitamin D (hypercalcaemia and renal stones).

### Risk of Bias Assessment for Individual Studies

We used the Cochrane Collaboration Risk of Bias tool^51^ to assess the following variables: sequence generation, allocation concealment, blinding of participants, personnel and outcome assessors, completeness of outcome data, evidence of selective outcome reporting and other potential threats to validity. Study quality was assessed independently by two investigators (ARM and DAJ), except for the five trials for which DAJ and/or ARM were investigators, which were assessed by CAC. Discrepancies were resolved by consensus.

### Definition of outcomes

The primary outcome of the meta-analysis was the proportion of participants experiencing one or more ARIs, with the definition of ARI encompassing events classified as URI, LRI and ARI of unclassified location (i.e. infection of the upper and/or lower respiratory tract). Secondary outcomes were incidence of URI and LRI, analysed separately; incidence of Emergency Department attendance and/or hospital admission for ARI; death due to ARI or respiratory failure; use of antibiotics to treat an ARI; absence from work or school due to ARI; incidence of serious adverse events; death due to any cause; and incidence of potential adverse reactions to vitamin D (hypercalcaemia and renal stones).

### Synthesis Methods

Data were analysed by DAJ; results were checked and verified by JDS. Our meta-analysis approach followed published guidelines.^52^ The primary comparison was of participants randomised to vitamin D vs. placebo: this was performed for all of the outcomes listed above. For trials that included higher-dose, lower-dose and placebo arms, data from higher-dose and lower-dose arms were pooled for analysis of the primary comparison. A secondary comparison of participants randomised to higher vs. lower doses of vitamin D was performed for the primary outcome only. A log odds ratio and its standard error was calculated for each outcome within each trial from the proportion of participants experiencing one or more events in the intervention vs. control arm. These were meta-analysed in a random effects model using the Metan package^53^ within STATA IC v14.2 to obtain a pooled odds ratio with a 95% confidence interval and a measure of heterogeneity summarized by the I^2^ statistic and its corresponding P value.

### Exploration of variation in effects

To explore reasons for heterogeneity of effect of the intervention between trials we performed a stratified analysis according to baseline vitamin D status (serum 25[OH]D <25 *vs*. 25-49.9 *vs*. 50-74.9 *vs*. ≥75 nmol/L) and according to age at baseline (<1 *vs*. 1-15.99 *vs*. 16-64.99 *vs*. ≥65 yrs). We additionally conducted sub-group analyses according to vitamin D dosing regimen (administration of daily *vs*. weekly *vs*. monthly or less frequent doses), dose size (daily equivalent <400 IU *vs*. 400-1000 IU *vs*. 1001-2000 IU *vs*. >2,000 IU), trial duration (≤12 months *vs*. >12 months) and presence of airway disease (trial restricted to participants with asthma *vs*. those restricted to participants with COPD *vs*. those in which participants without airway disease were eligible). The thresholds for baseline 25(OH)D concentration used in sub-group analyses were selected *a priori* on the basis that they represent cut-offs that are commonly used to distinguish profound vitamin D deficiency (<25 nmol/L), moderate vitamin D deficiency (25-49.9 nmol/L) and sub-optimal vitamin D status (50-74.9 nmol/L).^54^ An exploratory analysis restricted to studies with optimal dosing frequency, dose size and duration was also performed.

To investigate factors associated with heterogeneity of effect between subgroups of trials, we performed multivariable meta-regression analysis on trial-level characteristics, namely, dose frequency, dose size and trial duration, to produce an adjusted odds ratio, a 95% confidence interval and a P value for interaction for each factor. Independent variables were dichotomised to create a more parsimonious model (serum 25(OH)D of <25 vs. ≥25 nmol/L; administration of daily vs. non-daily doses; daily equivalent of ≤1000 IU vs. >1000 IU; and trial duration of ≤12 vs. >12 months). The meta-regression analysis excluded data from two trials that included higher-dose, lower-dose and placebo arms,^18,47^ since the higher-dose and lower-dose arms spanned the 1,000 IU/day cut-off, rendering them unclassifiable for the purposes of this analysis.

### Quality Assessment Across Studies

For the primary analysis, the likelihood of publication bias was investigated through the construction of a contour-enhanced funnel plot.^55^ We used the five GRADE considerations (study limitations, consistency of effect, imprecision, indirectness and publication bias)^56^ to assess the quality of the body of evidence contributing to analyses of the primary efficacy outcome and major secondary outcomes of our meta-analysis.

### Sensitivity analyses

We conducted three exploratory sensitivity analyses for the primary comparison of the primary outcome: one excluded RCTs where risk of bias was assessed as being unclear; one excluded RCTs in which incidence of ARI was not the primary or co-primary outcome; and one substituted diary-defined ARI events (available for 2598 participants) for survey-defined ARI events (available for n=16,000 participants) from the trial by Pham et al.^44^

## Results

### Study selection and data obtained

The study selection process is illustrated in Figure 1. Our search identified a total of 1,528 unique studies that were assessed for eligibility, of which 45 studies with a total of 73,384 randomised participants fulfilled eligibility criteria. Studies for which full text was reviewed prior to exclusion due to ineligibility are listed in Table S1. Of the 45 eligible studies identified, 34 compared effects of a single vitamin D regimen vs. placebo only,^5-17,19,20,22,23,25-28,31,33,36,38,39,41-45,48-50^ 5 compared effects of higher-dose, lower-dose and placebo arms,^18,21,24,40,47^ and 6 compared effects of higher-vs. lower-dose regimens of vitamin D only.^29,32,34,35,37,46^ Stratified aggregate data were sought and obtained for all but 3 eligible studies.^48-50^ Data for the primary outcome (proportion of participants with one or more ARI) were obtained for 46,331 (98.0%) of 47,262 participants in 42 studies.^5-17,31-47^

**Figure 1:**
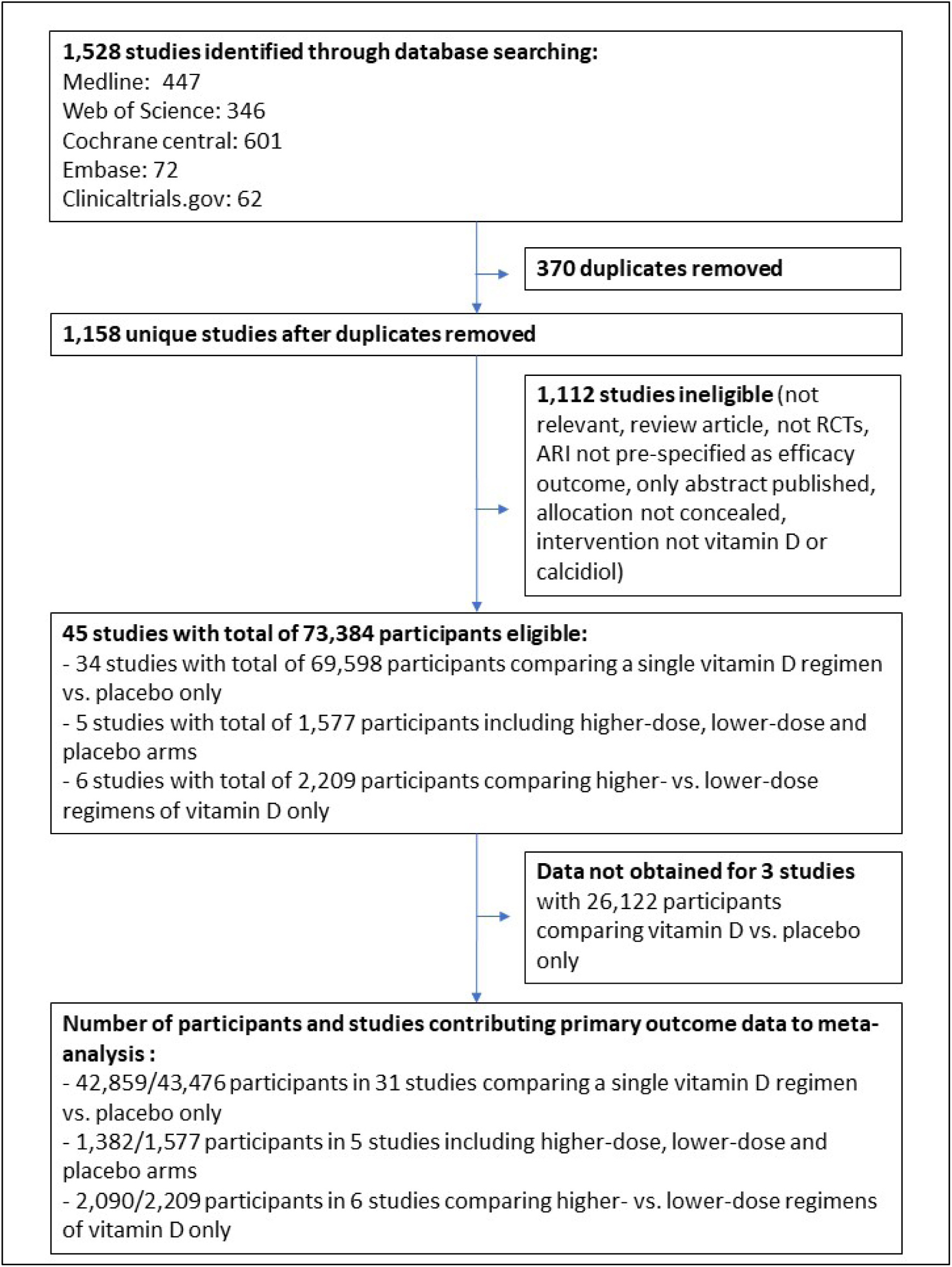
Flow chart of study selection

### Study and participant characteristics

Characteristics of the 42 studies contributing data to this meta-analysis and their participants are presented in Table 1. Trials were conducted in 18 different countries on 5 continents, and enrolled participants of both sexes from birth to 95 years of age. Baseline serum 25(OH)D concentrations were determined in 34 of 42 trials: mean baseline 25(OH)D concentration ranged from 18.9 to 90.9 nmol/L (to convert to ng/ml, divide by 2.496). Forty-one studies administered oral vitamin D3 to participants in the intervention arm, while 1 study administered oral 25(OH)D. Vitamin D was given as monthly to 3-monthly bolus doses in 13 studies; as weekly doses in 6 studies; as daily doses in 21 studies; and as a combination of bolus and daily doses in 2 studies. Trial duration ranged from 8 weeks to 5 years. Incidence of ARI was primary or co-primary outcome for 22 studies, and a secondary outcome for 20 studies.

**Table 1:**
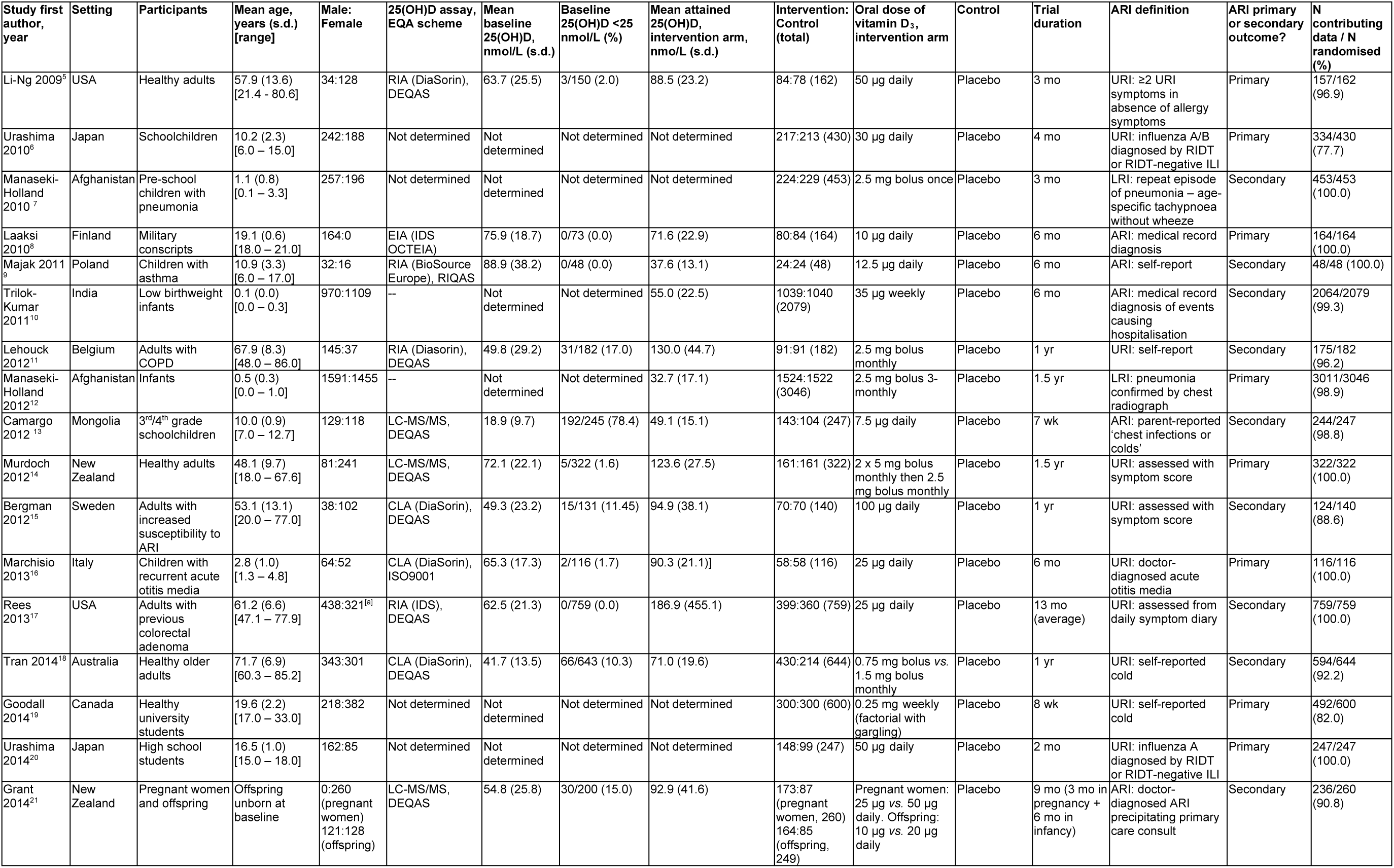
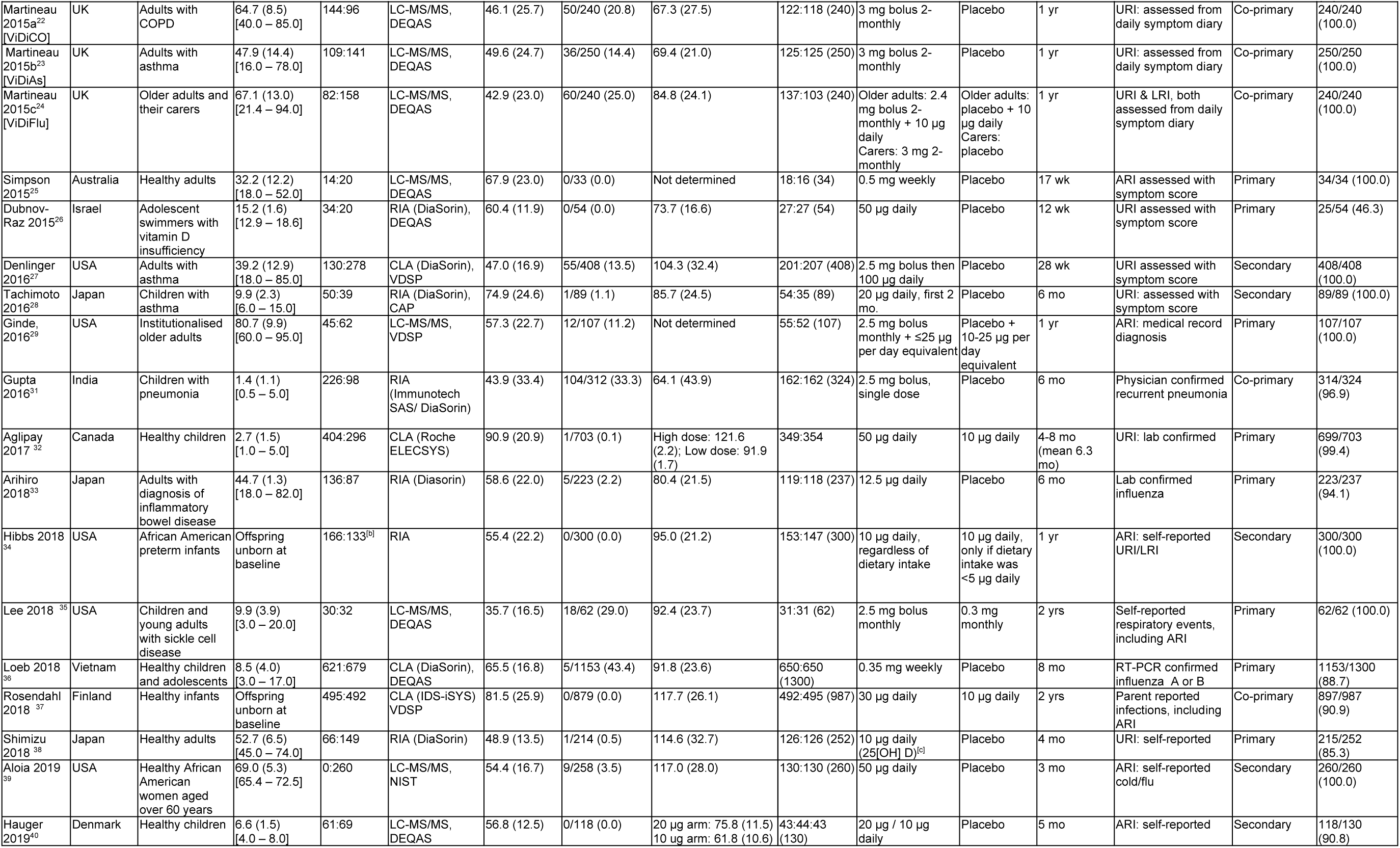
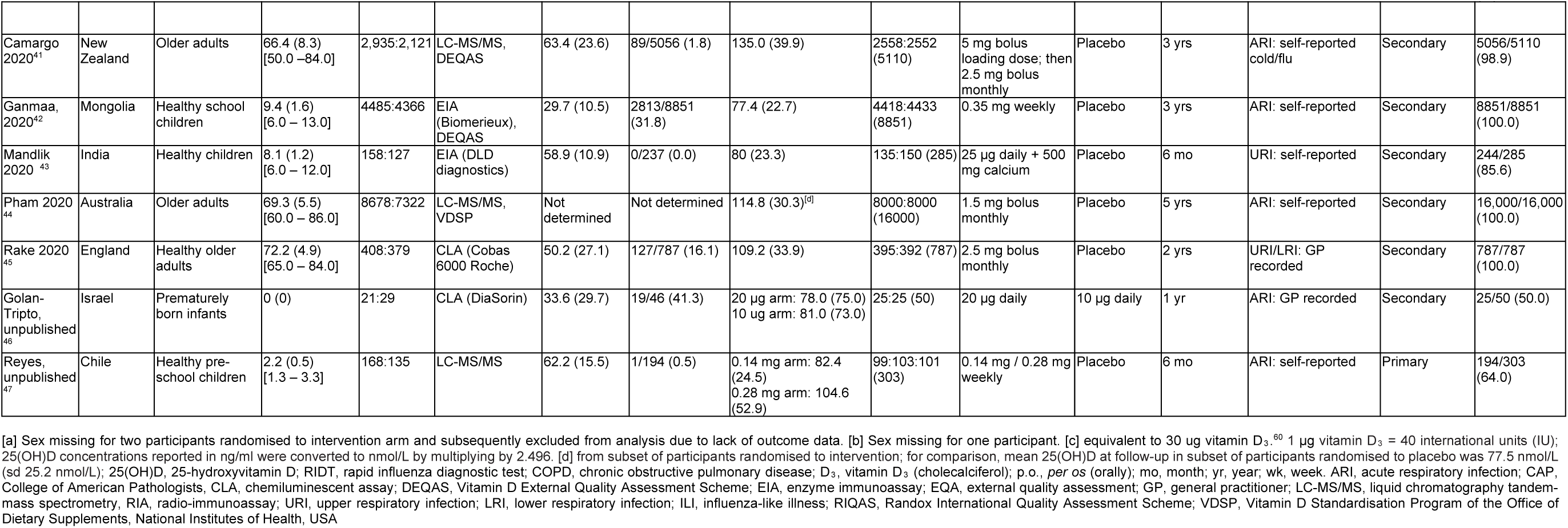
Characteristics of the 42 eligible trials and their participants

### Risk of Bias Within Studies

Details of the risk of bias assessment are provided in supplementary Table S2. Four trials were assessed as being at unclear risk of bias due to high loss to follow-up. In the trial by Laaksi and colleagues,^8^ 37% of randomised participants were lost to follow-up. In the trial by Dubnov-Raz and colleagues,^26^ 52% of participants did not complete all symptom questionnaires. In the unpublished trial by Reyes and colleagues,^47^ loss to follow-up ranged from 33% to 37% across the three study arms, and in the unpublished trial by Golan-Tripto and colleagues,^46^ 50% of participants were lost to follow-up. All other trials were assessed as being at low risk of bias for all seven aspects assessed.

### Overall Results, Primary Outcome

For the primary comparison of vitamin D vs. placebo control, supplementation resulted in a statistically significant reduction in the proportion of participants experiencing at least one ARI (Odds Ratio [OR] 0.91, 95% Confidence Interval [CI] 0.84 to 0.99; 44,009 participants in 36 studies; Figure 2, Table 2; Cates Plot, Figure S1). Heterogeneity of effect was moderate (I^2^ 37.2%, P for heterogeneity 0.01).

**Table 2:** Placebo controlled RCTs: Proportion of participants experiencing at least one acute respiratory infection, overall and stratified by potential effect-modifiers

| Potential effect-modifier | No of trials | Proportion with ≥1 ARI, intervention group (%) | Proportion with ≥1 ARI, control group (%) | Odds ratio (95% CI) | I<sup>2</sup> % | P for heterogeneity |
| --- | --- | --- | --- | --- | --- | --- |
| **Overall** | 36 | 13685/22288 (61.4) | 13565/21721 (62.5) | 0.91 (0.84 to 0.99) | 37.2 | 0.01 |
| **Baseline 25(OH)D, nmol/L<sup>[a]</sup>** |  |  |  |  |  |  |
| <25 | 19 | 1348/1798 (75.0) | 1388/1819 (76.3) | 0.78 (0.53 to 1.16) | 47.2 | 0.01 |
| 25 – 49.9 | 28 | 3439/4666 (73.7) | 3347/4501 (74.4) | 1.03 (0.92 to 1.15) | 0.4 | 0.46 |
| 50 – 74.9 | 29 | 1679/2839 (59.1) | 1565/2578 (60.7) | 0.90 (0.76 to 1.06) | 11.2 | 0.29 |
| ≥75 | 25 | 945/1543 (61.2) | 908/1471 (61.7) | 0.97 (0.81 to 1.16) | 0.0 | 0.78 |
| **Dosing frequency** |  |  |  |  |  |  |
| Daily | 18 | 1056/2134 (49.5) | 1020/1871 (54.5) | 0.75 (0.61 to 0.93) | 52.5 | 0.005 |
| Weekly | 6 | 4482/6421 (69.8) | 4447/6335 (70.2) | 0.97 (0.88 to 1.06) | 0.0 | 0.48 |
| Monthly or less frequently | 12 | 8147/13733 (59.3) | 8098/13515 (59.9) | 0.98 (0.93 to 1.03) | 0.0 | 0.57 |
| **Daily dose equivalent, IU<sup>[b]</sup>** |  |  |  |  |  |  |
| <400 | 2 | 482/1175 (41.0) | 511/1133 (45.1) | 0.65 (0.31 to 1.37) | 86.3 | 0.007 |
| 400-1000 | 10 | 656/1236 (53.1) | 627/1069 (58.7) | 0.70 (0.55 to 0.89) | 31.2 | 0.16 |
| 1001-2000 | 15 | 9946/15885 (62.6) | 10022/15817 (63.4) | 0.97 (0.92 to 1.03) | 1.0 | 0.44 |
| >2000 | 7 | 2291/3462 (66.2) | 2250/3444 (65.3) | 1.05 (0.84 to 1.31) | 37.1 | 0.15 |
| **Trial duration, months** |  |  |  |  |  |  |
| ≤12 | 29 | 1977/4887 (40.5) | 1866/4368 (42.7) | 0.82 (0.72 to 0.93) | 38.1 | 0.02 |
| >12 | 7 | 11708/17401 (67.3) | 11699/17353 (67.4) | 0.99 (0.95 to 1.04) | 0.0 | 0.91 |
| **Age, yrs<sup>[a]</sup>** |  |  |  |  |  |  |
| <1 | 5 | 875/2901 (30.2) | 839/2796 (30.0) | 0.95 (0.82 to 1.10) | 18.7 | 0.30 |
| 1-15.9 | 15 | 4297/5994 (71.7) | 4303/5877 (73.2) | 0.71 (0.57 to 0.90) | 46.0 | 0.03 |
| 16-64.9 | 21 | 3137/4876 (64.3) | 3087/4727 (65.3) | 0.97 (0.93 to 1.09) | 11.5 | 0.31 |
| ≥65 | 16 | 5376/8589 (62.6) | 5352/8394 (63.4) | 0.96 (0.90 to 1.02) | 0.0 | 0.67 |
| **Airway disease** |  |  |  |  |  |  |
| Asthma only | 4 | 203/404 (50.2) | 202/391 (51.7) | 0.73 (0.36 to 1.49) | 71.7 | 0.01 |
| COPD only | 2 | 106/208 (51.0) | 104/207 (50.2) | 1.01 (0.68 to 1.51) | 0.0 | 0.71 |
| Unrestricted | 30 | 13376/21676 (61.7) | 13259/21123 (62.8) | 0.91 (0.84 to 0.99) | 35.0 | 0.03 |

[a] The number of trials in each category for this variable adds up to more than 36, since this is a participant-level variable, i.e. some trials contributed data from participants who fell into more than one category
[b] Data from two trials that included higher-dose, lower-dose and placebo arms<sup>18,47</sup> are excluded from this sub-group analysis, since the higher-dose and lower-dose arms spanned the 1,000 IU/day cut-off, rendering them unclassifiable

**Figure 2:**
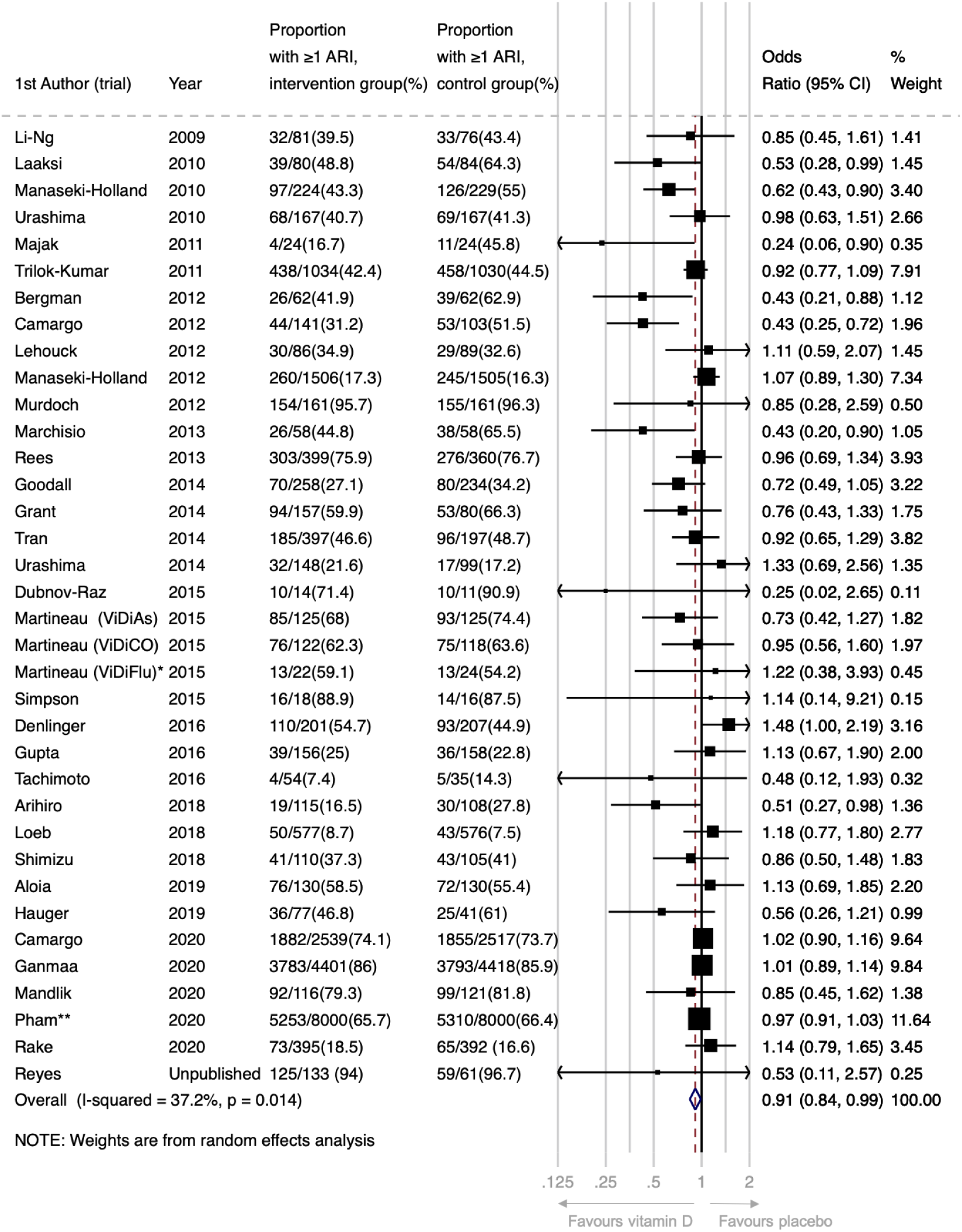
Forest plot of placebo-controlled RCTs reporting proportion of participants experiencing 1 or more acute respiratory infection. *This analysis includes data from the subset of ViDiFlu trial participants who were randomised to vitamin D vs. placebo control. **For this trial, participants were asked to report the occurrence of ARI during the one month prior to completing each annual urvey (max surveys=5). The numerator is the number of participants who reported an ARI on at least one survey. The ARI outcomes for participants who completed fewer than 5 surveys and who did not report an ARI (N=2239; 14%) were estimated based on the % affected among those who completed all 5 surveys (N=12152; 76%).

For the secondary comparison of higher-vs. lower-dose vitamin D, we observed no statistically significant difference in the proportion of participants with at least one ARI (OR 0.87, 95% CI 0.73 to 1.04; 3,047 participants in 11 studies; I^2^ 0.0%, P for heterogeneity 0.50; Figure S2).

### Sub-group analyses, Primary Outcome

To investigate reasons for the observed heterogeneity of effect for the primary comparison of vitamin D vs. placebo control, we stratified this analysis by two participant-level factors (baseline vitamin D status and age) and by four trial-level factors (dose frequency, dose size, trial duration, and airway disease comorbidity). Results are presented in Table 2 and Figures S3-S8. No statistically significant effect of vitamin D was seen for participants with baseline 25(OH)D <25 nmol/L (OR 0.78, 95% CI 0.53 to 1.16; 3,617 participants in 19 studies), 25-49.9 nmol/L (OR 1.03, 95% CI 0.92 to 1.15; 9,167 participants in 28 studies), 50-74.9 nmol (OR 0.90, 95% CI 0.76 to 1.06; 5,417 participants in 29 studies), or ≥75 nmol/L (OR 0.97, 95% CI 0.81 to 1.16; 3,014 participants in 25 studies; Figure S3). A statistically significant protective effect of vitamin D was seen for participants aged 1-15.9 years (OR 0.71, 95% CI 0.57 to 0.90; 11,871 participants in 15 studies), but not in participants aged <1 year (OR 0.95, 95% CI 0.82 to 1.10; 5,697 participants in 5 studies), 16-64.99 years (OR 0.97, 95% CI 0.93 to 1.09; 9,603 participants in 21 studies), or ≥65 years (OR 0.96, 95% CI 0.90 to 1.02; 16,983 participants in 16 studies; Figure S7).

With regard to dosing frequency, a statistically significant protective effect was seen for trials where vitamin D was given daily (OR 0.75, 95% CI 0.61 to 0.93; 4,005 participants in 18 studies), but not for trials in which it was given weekly (OR 0.97, 95% CI 0.88 to 1.06; 12,756 participants in 6 studies), or monthly to 3-monthly (OR 0.98, 95% CI 0.93 to 1.03; 27,248 participants in 12 studies; Figure S4). Statistically significant protective effects of the intervention were also seen in trials where vitamin D was administered at daily equivalent doses of 400-1000 IU (OR 0.70, 95% CI 0.55 to 0.89; 2,305 participants in 10 studies), but not where the daily dose equivalent was <400 IU (OR 0.65, 95% CI 0.31 to 1.37; 2,308 participants in 2 studies), 1001-2000 IU (OR 0.97, 95% CI 0.92 to 1.03; 31,702 participants in 15 studies), or >2000 IU (OR 1.05, 95% CI 0.84 to 1.31; 6,906 participants in 7 studies; Figure S5). Statistically significant protective effects were also seen for trials with a duration of ≤12 months (OR 0.82, 95% CI 0.72 to 0.93; 9,255 participants in 29 studies) but not in those lasting >12 months (OR 0.99, 95% CI 0.95 to 1.04; 34,754 participants in 7 studies; Figure S6).

Finally, statistically significant protective effects were also seen for trials that were not restricted to participants with asthma or COPD (OR 0.91, 95% CI 0.84 to 0.99; 42,799 participants in 30 studies), but not in trials that exclusively enrolled participants with asthma (OR 0.73, 95% CI 0.36 to 1.49; 795 participants in 4 studies), or COPD (OR 1.01, 95% 0.68 to 1.51; 415 participants in 2 studies; Figure S8).

An exploratory analysis restricted to eight placebo-controlled trials investigating effects of daily dosing at doses of 400-1000 IU/day with duration ≤12 months (for which mean baseline 25(OH)D level ranged from 54.8 nmol/L to 88.9 nmol/L) showed a statistically significant reduction in the proportion of participants experiencing at least one ARI (OR 0.58, 95% CI 0.45 to 0.75; 1,232 participants in 8 studies; Figure S9; Cates Plot, Figure S1). Heterogeneity of effect for this exploratory analysis was low (I^2^ 0.0%, P for heterogeneity 0.67).

### Multivariable Meta-Regression Analysis

Multivariable meta-regression analysis of trial-level sub-groups did not identify a statistically significant interaction between allocation to vitamin D vs. placebo and dose frequency, size or trial duration (Table S3).

### Secondary outcomes

Meta-analysis of secondary outcomes was performed for results of placebo-controlled trials only; results are presented in Table 3. Overall, without consideration of participant-or trial-level factors, vitamin D supplementation did not have a statistically significant effect on the proportion of participants with one or more URI, LRI, courses of antimicrobials for ARI, work/school absences due to ARI, hospitalisations or emergency department attendances for ARI, serious adverse events of any cause, death due to ARI or respiratory failure, death due to any cause, or episodes of hypercalcaemia or renal stones.

**Table 3:** Placebo-controlled studies: Secondary outcomes

| Variables | No of trials | Proportion with $\geq 1$ event, intervention group (%) | Proportion with $\geq 1$ event, control group (%) | Odds ratio (95% CI) | I<sup>2</sup> % | P for heterogeneity |
| --- | --- | --- | --- | --- | --- | --- |
| **Efficacy outcomes** |  |  |  |  |  |  |
| Upper respiratory infection* | 28 | 7931/13493 (58.8) | 7823/13034 (60.0) | 0.96 (0.90 to 1.02) | 4.4 | 0.40 |
| Lower respiratory infection* | 14 | 3583/12167 (29.4) | 3601/12027 (29.9) | 0.98 (0.93 to 1.04) | 0.0 | 0.56 |
| Emergency department attendance and/or hospital admission due to ARI | 18 | 117/9887 (1.2) | 125/9769 (1.3) | 0.90 (0.70 to 1.16) | 0.0 | 1.00 |
| Death due to ARI or respiratory failure | 33 | 14/13612 (0.1) | 11/13058 (0.1) | 1.04 (0.61 to 1.78) | 0.0 | 1.00 |
| Use of antibiotics to treat an ARI* | 13 | 2035/7562 (26.9) | 2082/7423 (28.0) | 0.91 (0.82 to 1.02) | 13.4 | 0.31 |
| Absence from work or school due to ARI | 10 | 378/1527 (24.7) | 364/1044 (34.9) | 0.91 (0.69 to 1.20) | 35.3 | 0.13 |
| **Safety outcomes** |  |  |  |  |  |  |
| Serious adverse event of any cause* | 35 | 545/13861 (3.9) | 554/13326 (4.2) | 0.97 (0.86 to 1.09) | 0.0 | 1.00 |
| Death due to any cause | 34 | 117/13854 (0.8) | 97/13293 (0.7) | 1.15 (0.90 to 1.49) | 0.0 | 1.00 |
| Hypercalcaemia | 21 | 50/9294 (0.5) | 39/8919 (0.4) | 1.20 (0.81 to 1.79) | 0.0 | 1.00 |
| Renal stones | 20 | 110/11540 (1.0) | 128/11138 (1.1) | 0.85 (0.66 to 1.10) | 0.0 | 1.00 |

\* This analysis includes a subset of participants in the trial by Pham et al, who completed symptom diaries.

### Risk of bias across studies

A funnel plot for the proportion of participants experiencing at least one ARI (Figure S10) showed left-sided asymmetry, confirmed with an Egger’s regression test^57^ (P=0.008). This might reflect heterogeneity of effect across trials, or publication bias arising from omission of small trials showing non-protective effects of vitamin D from the meta-analysis.^58^ Given the latter possibility, the quality of the body of evidence contributing to analyses of the primary efficacy outcome and major secondary outcomes was downgraded to moderate (Table S4).

### Sensitivity Analyses

Results of exploratory sensitivity analyses are presented in Table S5. Meta-analysis of the proportion of participants in placebo-controlled trials experiencing at least one ARI, excluding 4 studies assessed as being at unclear risk of bias,^8,26,46,47^ revealed protective effects of vitamin D supplementation consistent with the main analysis (OR 0.93, 95% CI 0.86 to 1.00; 43,626 participants in 33 studies). Sensitivity analysis for the same outcome, excluding 18 placebo-controlled trials that investigated ARI as a secondary outcome, did not show a statistically significant protective effect (OR 0.89, 95% CI 0.77 to 1.03; 7,537 participants in 18 studies). A sensitivity analysis for the same outcome, substituting diary-defined ARI events (available for 2598 participants) for survey-defined ARI events (available for n=16,000 participants) in the trial by Pham et al^44^ revealed protective effects of vitamin D supplementation consistent with the main analysis (OR 0.90, 95% CI 0.83 to 0.99; 30,607 participants in 36 studies).

## Discussion

This updated meta-analysis of RCTs of vitamin D supplementation for the prevention of ARI includes data from an additional 17 studies completed since December 2015, when we performed the final literature search for our prior individual participant data meta-analysis.^30^ For expediency during the COVID-19 pandemic, we used a trial-level approach for this update, which includes data from a total of 46,331 participants in 42 trials. Overall, we report a modest statistically significant protective effect of vitamin D supplementation, as compared with placebo (OR 0.91, 95% CI 0.84 to 0.99). As expected, there was significant heterogeneity (P=0.01) across trials, which might have led to an under-estimate of the protective effect, and contributed to the asymmetry observed in the funnel plot.^58^ Alternatively, left-sided asymmetry in the funnel plot may reflect publication bias, which might have led to an over-estimate of the protective effect. In contrast to findings of our previous meta-analysis,^30^ we did not observed enhanced protection in those with the lowest 25(OH)D levels at baseline. However, there was evidence that efficacy of vitamin D supplementation varied according to dosing regimen and trial duration, with protective effects associated with daily administration of doses of 400-1000 IU vitamin D given for ≤12 months. An exploratory analysis restricted to data from 8 trials fulfilling these design criteria revealed a larger protective effect (OR 0.58, 95% CI 0.45 to 0.75) without significant heterogeneity across trials (P for heterogeneity 0.67).

The magnitude of the overall protective effect seen in the current analysis (OR 0.91, 95% CI 0.84 to 0.99) is modest, and similar to the value reported in our previous meta-analysis of individual participant data (adjusted OR 0.88, 95% CI 0.81 to 0.96).^30^ In keeping with our previous study, the point estimate for this effect was lower among those with baseline 25(OH)D <25 nmol/L than in those with higher baseline vitamin D status. However, in contrast to our previous finding, a statistically significant protective effect of vitamin D was not seen in those with the lowest 25(OH)D concentrations. This difference reflects the inclusion of null data from four new RCTs in which vitamin D was given in relatively high doses at weekly or monthly intervals over 2-5 years. ^41,42,44,45^ Null results of these studies contrast with protective effects reported from earlier trials in which smaller daily doses of vitamin D were given over shorter periods. ^8,9,13,16^ These differing findings suggest that the frequency, amount and duration of vitamin D supplementation may be key determinants of its protective efficacy. In keeping with this hypothesis, statistically significant protective effects of vitamin D were seen for meta-analysis of trials where vitamin D was given daily; where it was given at doses of 400-1000 IU/day; and where it was given for 12 months or less. When results of trials that investigated daily administration of 400-1000 IU over ≤12 months were pooled in an exploratory meta-analysis, a protective effect was seen (OR 0.58, 95% CI 0.45 to 0.75) with low heterogeneity (I^2^ 0.0%, P for heterogeneity 0.67). Greater protective efficacy of lower vs higher doses of vitamin D might reflect deleterious effects of higher-dose vitamin D on its own metabolism, or on host responses to respiratory pathogens: head-to-head mechanistic studies in individuals randomised to different regimens of vitamin D supplementation are needed to investigate this issue.

The current study has several strengths: it contains the very latest RCT data available in this fast-moving field, including findings from three large phase 3 trials published in 2020^41,42,44^ as well as some as-yet unpublished studies.^46,47^ The inclusion of additional studies allowed us to analyse results of placebo-controlled studies vs. high-dose / low-dose studies separately, and gave us the power to investigate reasons for heterogeneity of effect observed across trials. For example, we could distinguish the effects of daily vs. weekly dosing, which were previously pooled.^30^

Our work also has limitations. Given the need to generate a rapid update of our previous work in the context of the COVID-19 pandemic, we meta-analysed aggregate (trial-level) data, rather than individual participant data; this allowed us to proceed rapidly, without the delays introduced by the need to establish multiple data sharing agreements. However, we did contact authors to get unpublished estimates of effect that were stratified by pre-defined baseline 25(OH)D levels, harmonised across studies: thus, we were able to provide accurate data for the major participant-level effect-modifier of interest. Despite the large number of trials overall, relatively few compared effects of lower-vs. higher-dose vitamin D: our power for this secondary comparison was therefore limited. We lacked the individual participant data to investigate race/ethnicity and obesity as potential effect-modifiers. We also could not account for other factors that might influence the efficacy of vitamin D supplements for ARI prevention (e.g., taking the supplement with or without food) or secular trends that would influence trials, such as the increased societal use of vitamin D supplements;^59^ concurrent use of standard dose vitamin D supplements or multivitamins in the “placebo” group would effectively render these as high-vs. low-dose trials and potentially drive results toward the null. A final limitation relates to the funnel plot, which suggests that the overall effect size may have been over-estimated due to publication bias; we have mitigated this by inclusion of data from unpublished studies identified by searching clinicaltrials.gov where this was obtainable.

In summary, this updated meta-analysis of data from RCTs of vitamin D for the prevention of ARI showed a statistically significant overall protective effect of the intervention. The protective effect was heterogenous across trials; it also may have been over-estimated due to publication bias. In contrast to findings of our previous meta-analysis of individual participant data, we did not see a protective effect of vitamin D supplementation among those with the lowest baseline vitamin D status. The vitamin D dosing regimen of most benefit was daily and used standard doses (e.g., 400 to 1000 IU) for up to 12 months. The relevance of these findings to COVID-19 is not known and requires investigation.

## Data Availability

The study dataset is available from.

## Acknowledgements

This study was conducted without external funding. DAJ is supported by a Barts Charity Lectureship (ref MGU045). ARM is supported by the United Kingdom Office for Students. The views expressed are those of the authors and not necessarily those of Barts Charity or the Office for Students. Sources of support for individual trials are detailed in Supplementary Material. We thank all the people who participated in primary randomised controlled trials, and the teams who conducted them.

## Author Contributions

DAJ and ARM wrote the study protocol and designed statistical analyses. DAJ, CAC and ARM assessed eligibility of studies for inclusion and performed risk of bias assessments. Statistical analyses were done by DAJ; results were checked and verified by JDS. DAJ and ARM wrote the first draft of the report. All authors revised it critically for important intellectual content, gave final approval of the version to be published, and agreed to be accountable for all aspects of the work in ensuring that questions related to the accuracy or integrity of any part of the work were appropriately investigated and resolved.

## Transparency Declaration

DAJ and ARM are the manuscript’s guarantors and they affirm that this is an honest, accurate, and transparent account of the study being reported and that no important aspects of the study have been omitted. All analyses were pre-specified in the study protocol, other than the exploratory analyses whose results are presented in Table 2 (sub-group analyses by age and presence of asthma/COPD, requested by reviewers), Table S5 and Figure S7.

## Data Sharing

the study dataset is available from.

## Supplementary Material

### Methods

#### Search Strategies

##### A. Medline

###### Cochrane Highly Sensitive Search Strategy for identifying randomised controlled trials

#1. randomized controlled trial [pt] OR controlled clinical trial [pt] OR randomized [tiab] OR placebo [tiab] OR drug therapy [sh] OR randomly [tiab] OR trial [tiab] OR groups [tiab]

#2. animals [mh] NOT humans [mh]

#3. #1 NOT #2

###### Terms specific to vitamin D

#4. Vitamin D OR vitamin D2 OR vitamin D3 OR cholecalciferol OR ergocalciferol OR alphacalcidol OR alfacalcidol OR calcitriol OR paricalcitol OR doxerocalciferol

###### Terms specific to acute respiratory infection

#5. Acute Respiratory Infection OR Upper Respiratory Infection OR Lower Respiratory Infection OR Respiratory Tract Infection OR Common Cold OR Sinusitis OR Pharyngitis OR Laryngitis OR Laryngotracheobronchitis OR Tonsillitis OR peritonsillar abscess OR Croup OR Epiglottitis OR supraglottitis OR Otitis Media OR Pneumonia OR Bronchopneumonia OR Bronchitis OR Pleurisy OR Pleuritis

###### Combination of terms to identify randomised controlled trials of vitamin D for the prevention of acute respiratory infection

#3 AND #4 AND #5

##### B. EMBASE

###### Terms for identifying randomised controlled trials

#1 ‘randomized controlled trial’/exp OR ‘single blind procedure’/exp OR ‘double blind procedure’/exp OR ‘crossover procedure’/exp

#2 random*:ab,ti OR placebo*:ab,ti OR crossover*:ab,ti OR ‘cross over’:ab,ti OR allocat*:ab,ti OR ((singl* OR doubl*) NEXT/1 blind*):ab,ti OR trial:ti

#3. #1 OR #2

###### Terms specific to vitamin D

#4. vitamin AND d OR vitamin AND d2 OR vitamin AND d3 OR cholecalciferol OR ergocalciferol OR alphacalcidol OR alfacalcidol OR calcitriol OR paricalcitol OR doxerocalciferol

###### Terms specific to acute respiratory infection

#5. acute AND respiratory AND infection OR upper AND respiratory AND infection OR lower AND respiratory AND infection OR respiratory AND tract AND infection OR common AND cold OR sinusitis OR pharyngitis OR laryngitis OR laryngotracheobronchitis OR tonsillitis OR peritonsillar AND abscess OR croup OR epiglottitis OR supraglottitis OR otitis AND media OR pneumonia OR bronchopneumonia OR bronchitis OR pleurisy OR pleuritis

###### Combination of terms to identify randomised controlled trials of vitamin D for the prevention of acute respiratory infection

#3 AND #4 AND #5

##### C. Cochrane Central

###### Terms specific to vitamin D

#1. Vitamin D OR vitamin D2 OR vitamin D3 OR cholecalciferol OR ergocalciferol OR alphacalcidol OR alfacalcidol OR calcitriol OR paricalcitol OR doxerocalciferol

###### Terms specific to acute respiratory infection

#2. Acute Respiratory Infection OR Upper Respiratory Infection OR Lower Respiratory Infection OR Respiratory Tract Infection OR Common Cold OR Sinusitis OR Pharyngitis OR Laryngitis OR Laryngotracheobronchitis OR Tonsillitis OR peritonsillar abscess OR Croup OR Epiglottitis OR supraglottitis OR Otitis Media OR Pneumonia OR Bronchopneumonia OR Bronchitis OR Pleurisy OR Pleuritis

###### Combination of terms to identify randomised controlled trials of vitamin D for the prevention of acute respiratory infection

#1 AND #2

##### D. Web of Science

TS =(Vitamin D OR vitamin D2 OR vitamin D3 OR cholecalciferol OR ergocalciferol OR alphacalcidol OR alfacalcidol OR calcitriol OR paricalcitol OR doxerocalciferol) AND TS =(Acute Respiratory Infection OR Upper Respiratory Infection OR Lower Respiratory Infection OR Respiratory Tract Infection OR Common Cold OR Sinusitis OR Pharyngitis OR Laryngitis OR Laryngotracheobronchitis OR Tonsillitis OR peritonsillar abscess OR Croup OR Epiglottitis OR supraglottitis OR Otitis Media OR Pneumonia OR Bronchopneumonia OR Bronchitis OR Pleurisy OR Pleuritis) AND TS =(placebo* or random* or clinical trial* or double blind* or single blind* or rct)

##### E. ClinicalTrials.gov

Vitamin D AND respiratory AND infection

### Sources of support for participating trials

The trial by Aglipay and colleagues was supported by the competitive grants from the Canadian Institutes of Health Research Institutes of Human Development, Child and Youth Health and Nutrition, Metabolism and Diabetes (grant number MOP-114945) and the Thrasher Research Fund (award number 9113).

The trial by Aloia and colleagues was supported by the National Institute of Aging (grant number R01-AG032440-01A2).

The trial by Arihiro and colleagues was supported by the Ministry of Education, Culture, Sports, Science, and Technology in the Japan-Supported Program for the Strategic Research Foundation at Private Universities and funding from the Department of Gastroenterology and Hepatology, Jikei University of Medicine, Tokyo, Japan.

The trial by Bergman et al was supported by grants from the Swedish Research Council, the Strategic Research Foundation (SSF), the Swedish Heart and Lung foundation, Karolinska Institutet, Stockholm County Council and the Swedish Cancer Society as well as by the Magnus Bergwall and Åke Wiberg foundations.

The trials by Camargo and colleagues were supported by a grant for the Blue Sky Study from an anonymous foundation and the Massachusetts General Hospital; and the Health Research Council of New Zealand (grant number 10/400) and the Accident Compensation Corporation of New Zealand.

The trial by Ganmaa and colleagues was supported by the National Institutes of Health (Grant Number 1R01HL122624-01).

The trial by Ginde and colleagues was supported by NIH/NIA grant K23AG040708, NIH/NCATS Colorado CTSA Grant UL1TR001082, and the American Geriatrics Society Jahnigen Career Development Scholars Award.

The trial by Golan-Tripto and colleagues was supported by the Soroka JNF UK Clinical Research Scholar Program.

The trial by Goodall and colleagues was supported in part by the Canadian Institutes of Health Research [OPP 86940] and with in-kind support from Copan Italia, Bresica Italy.

The trial by Grant and colleagues was supported by the Health Research Council of New Zealand, Grant Number 09/215R.

The trial by Gupta and colleagues was supported by the Indian Council of Medical Research, New Delhi.

The trial by Hauger and colleagues was supported by Lundbeckfonden (grant number R180-2014-3481), by Brødrene Hartmann’s Fund (A26842), and by the European

Commission (FP7/2007–2013) under Grant Agreement 613977 for the ODIN Integrated Project (Food-based solutions for optimal vitamin D nutrition and health through the life cycle).

The trial by Hibbs and colleagues was supported by the National Heart, Lung, and Blood Institute and Office of Dietary Supplements (grant number R01HL109293).

The trial by Lee and colleagues was supported by the US Food and Drug Administration Orphan Product Development (grant number R01FD003894).

The trial by Loeb and colleagues was supported by the Institute for Infectious Diseases Research at McMaster University.

The trials by Manaseki-Holland and colleagues were supported by New Zealand Aid (ref: GRA/470/2) and The Wellcome Trust (ref: 082476).

The trial by Mandlik and colleagues was supported by a core grant from the Hirabai Cowasji Jehangir Medical Research Institute.

The trials by Martineau and colleagues were supported by the National Institute for Health Research under its Programme Grants for Applied Research Programme (Reference Number RP-PG-0407-10398).

The trial by Murdoch and colleagues was supported by the Health Research Council of New Zealand, grant number 09/302.

The trial by Pham and colleagues was supported by project grants from the National Health and Medical Research Council (grant numbers GNT1046681 and GNT1120682).

The trial by Reyes and colleagues was supported by Fondo Nacional de Investigacion y Desarrollo en Salud (ref. SA13I20173).

The trial by Rosendahl and colleagues was supported by the Foundation for Pediatric Research, the Finnish Medical Foundation, Governmental Subsidy for Clinical Research, the Päivikki and Sakari Sohlberg Foundation, the Academy of Finland, the Sigrid Jusélius Foundation, the Folkhälsan Research Foundation, the Novo Nordisk Foundation, the Orion Research Foundation, and Barncancerfonden.

The trial by Rake and colleagues was supported by the National Institute for Health Research Health Technology Assessment programme (ref. HTA 08/116/48).

The trial by Rees and colleagues was supported by the National Cancer Institute at the National Institutes of Health (grant numbers CA098286 and CA098286-S).

The trial by Shimizu and colleagues was supported by the FANCL Corporation.

The trial by Simpson and colleagues was supported by the Royal Hobart Hospital Research Foundation.

The trial by Tachimoto and colleagues was supported by the Ministry of Education, Culture, Sports, Science, and Technology in the Japan-Supported Program for the Strategic Research Foundation at Private Universities, JSPH KAKENHI Grant Number 23591553, and funding from Jikei University of Medicine, Tokyo, Japan.

The trial by Tran and colleagues was supported by the National Health and Medical Research Council of Australia, grant 613655.

The trial by Trilok Kumar and colleagues was supported by the Department of Biotechnology, Government of India (ref BT/PR-PR7489/PID/20/285/2006), Nutrition Third World and Sight and Life.

**Table S1:** Reasons for exclusion of potentially relevant studies.

| **First author, year** | **Reason for exclusion** |
| --- | --- |
| Somnath, 2017<sup>1</sup> | Ineligible: open-label trial |
| Jung, 2018<sup>2</sup> | Ineligible: ARI outcome not pre-specified |
| Ramos-Martínez, 2018<sup>3</sup> | Ineligible: intervention was administration of 1,25-dihydroxyvitamin D |
| Zhou, 2018<sup>4</sup> | Ineligible: open-label trial |
| Hueniken, 2019<sup>5</sup> | Ineligible: same trial as Aglipay et al<sup>6</sup> |
| Singh, 2019<sup>7</sup> | Ineligible: open-label trial |

**Table S2:** Risk of Bias Assessment

|  | Sequence generation | Allocation concealment | Blinding of participants and personnel | Blinding of outcome assessment | Incomplete outcome data | Selective reporting | Other bias |
| --- | --- | --- | --- | --- | --- | --- | --- |
| Li-Ng 2009<sup>8</sup> | ✓ | ✓ | ✓ | ✓ | ✓ | ✓ | ✓ |
| Urashima 2010<sup>9</sup> | ✓ | ✓ | ✓ | ✓ | ✓ | ✓ | ✓ |
| Manaseki-Holland 2010<sup>10</sup> | ✓ | ✓ | ✓ | ✓ | ✓ | ✓ | ✓ |
| Laaksi 2010<sup>11</sup> | ✓ | ✓ | ✓ | ✓ | ? | ✓ | ✓ |
| Majak 2011<sup>12</sup> | ✓ | ✓ | ✓ | ✓ | ✓ | ✓ | ✓ |
| Trilok-Kumar 2011<sup>13</sup> | ✓ | ✓ | ✓ | ✓ | ✓ | ✓ | ✓ |
| Lehouck 2012<sup>14</sup> | ✓ | ✓ | ✓ | ✓ | ✓ | ✓ | ✓ |
| Manaseki-Holland 2012<sup>15</sup> | ✓ | ✓ | ✓ | ✓ | ✓ | ✓ | ✓ |
| Camargo 2012<sup>16</sup> | ✓ | ✓ | ✓ | ✓ | ✓ | ✓ | ✓ |
| Murdoch 2012<sup>17</sup> | ✓ | ✓ | ✓ | ✓ | ✓ | ✓ | ✓ |
| Bergman 2012<sup>18</sup> | ✓ | ✓ | ✓ | ✓ | ✓ | ✓ | ✓ |
| Marchisio 2013<sup>19</sup> | ✓ | ✓ | ✓ | ✓ | ✓ | ✓ | ✓ |
| Rees 2013<sup>20</sup> | ✓ | ✓ | ✓ | ✓ | ✓ | ✓ | ✓ |
| Tran 2014<sup>21</sup> | ✓ | ✓ | ✓ | ✓ | ✓ | ✓ | ✓ |
| Goodall 2014<sup>22</sup> | ✓ | ✓ | ✓ | ✓ | ✓ | ✓ | ✓ |
| Urashima 2014<sup>23</sup> | ✓ | ✓ | ✓ | ✓ | ✓ | ✓ | ✓ |
| Grant 2014<sup>24</sup> | ✓ | ✓ | ✓ | ✓ | ✓ | ✓ | ✓ |
| Martineau 2015a<sup>25</sup> [ViDiCO] | ✓ | ✓ | ✓ | ✓ | ✓ | ✓ | ✓ |
| Martineau 2015b<sup>26</sup> [ViDiAs] | ✓ | ✓ | ✓ | ✓ | ✓ | ✓ | ✓ |
| Martineau 2015c<sup>27</sup> [ViDiFlu] | ✓ | ✓ | ✓ | ✓ | ✓ | ✓ | ✓ |
| Simpson 2015<sup>28</sup> | ✓ | ✓ | ✓ | ✓ | ✓ | ✓ | ✓ |
| Dubnov-Raz 2015<sup>29</sup> | ✓ | ✓ | ✓ | ✓ | ? | ✓ | ✓ |
| Denlinger 2016<sup>30</sup> | ✓ | ✓ | ✓ | ✓ | ✓ | ✓ | ✓ |
| Tachimoto 2016<sup>31</sup> | ✓ | ✓ | ✓ | ✓ | ✓ | ✓ | ✓ |
| Ginde 2016<sup>32</sup> | ✓ | ✓ | ✓ | ✓ | ✓ | ✓ | ✓ |
| Gupta 2016<sup>33</sup> | ✓ | ✓ | ✓ | ✓ | ✓ | ✓ | ✓ |
| Aglipay 2017<sup>6</sup> | ✓ | ✓ | ✓ | ✓ | ✓ | ✓ | ✓ |
| Arihiro 2018<sup>34</sup> | ✓ | ✓ | ✓ | ✓ | ✓ | ✓ | ✓ |
| Hibbs 2018<sup>35</sup> | ✓ | ✓ | ✓ | ✓ | ✓ | ✓ | ✓ |
| Lee 2018<sup>36</sup> | ✓ | ✓ | ✓ | ✓ | ✓ | ✓ | ✓ |
| Loeb 2018<sup>37</sup> | ✓ | ✓ | ✓ | ✓ | ✓ | ✓ | ✓ |
| Rosendahl 2018<sup>38</sup> | ✓ | ✓ | ✓ | ✓ | ✓ | ✓ | ✓ |
| Shimizu 2018<sup>39</sup> | ✓ | ✓ | ✓ | ✓ | ✓ | ✓ | ✓ |
| Aloia 2019<sup>40</sup> | ✓ | ✓ | ✓ | ✓ | ✓ | ✓ | ✓ |
| Hauger 2019<sup>41</sup> | ✓ | ✓ | ✓ | ✓ | ✓ | ✓ | ✓ |
| Camargo 2020<sup>42</sup> | ✓ | ✓ | ✓ | ✓ | ✓ | ✓ | ✓ |
| Ganmaa 2020<sup>43</sup> | ✓ | ✓ | ✓ | ✓ | ✓ | ✓ | ✓ |
| Mandlik 2020<sup>44</sup> | ✓ | ✓ | ✓ | ✓ | ✓ | ✓ | ✓ |
| Pham 2020<sup>45</sup> | ✓ | ✓ | ✓ | ✓ | ✓ | N/A | ✓ |
| Rake 2020<sup>46</sup> | ✓ | ✓ | ✓ | ✓ | ✓ | ✓ | ✓ |
| Golan-Tripto (unpublished)<sup>47</sup> | ✓ | ✓ | ✓ | ✓ | ? | N/A | ✓ |
| Reyes (unpublished)<sup>48</sup> | ✓ | ✓ | ✓ | ✓ | ? | N/A | ✓ |

✓ = low risk of bias; ? = unclear risk of bias N/A = not applicable (unpublished)

**Table S3:**
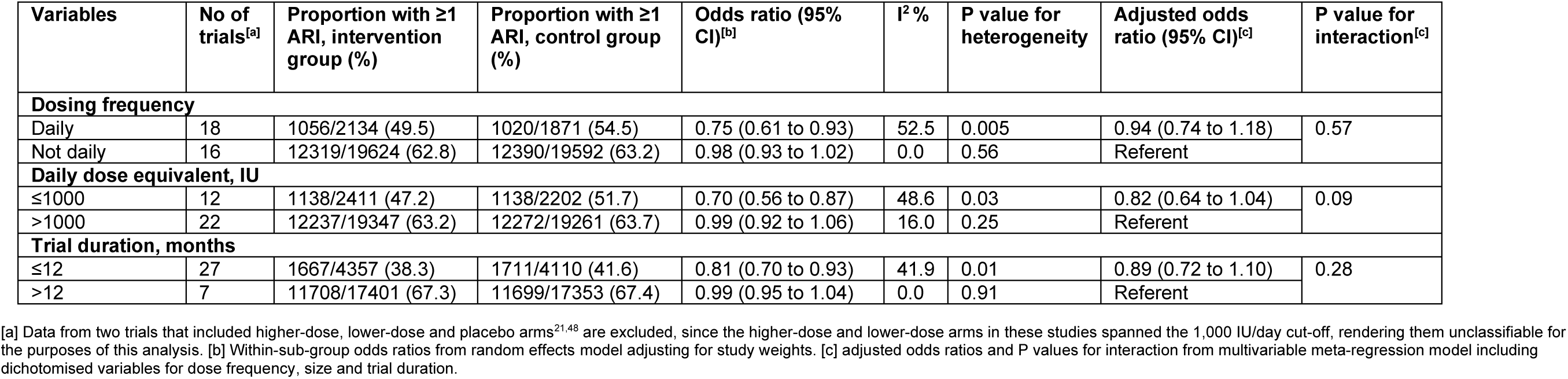
Multivariate meta-regression model for proportion of participants experiencing at least one acute respiratory tract infection, by trial-level subgroups.

| Variables | No of trials<sup>[a]</sup> | Proportion with ≥1 ARI, intervention group (%) | Proportion with ≥1 ARI, control group (%) | Odds ratio (95% CI)<sup>[b]</sup> | I<sup>2</sup> % | P value for heterogeneity | Adjusted odds ratio (95% CI)<sup>[c]</sup> | P value for interaction<sup>[c]</sup> |
| --- | --- | --- | --- | --- | --- | --- | --- | --- |
| Dosing frequency |  |  |  |  |  |  |  |  |
| Daily | 18 | 1056/2134 (49.5) | 1020/1871 (54.5) | 0.75 (0.61 to 0.93) | 52.5 | 0.005 | 0.94 (0.74 to 1.18) | 0.57 |
| Not daily | 16 | 12319/19624 (62.8) | 12390/19592 (63.2) | 0.98 (0.93 to 1.02) | 0.0 | 0.56 | Referent |  |
| Daily dose equivalent, IU |  |  |  |  |  |  |  |  |
| ≤1000 | 12 | 1138/2411 (47.2) | 1138/2202 (51.7) | 0.70 (0.56 to 0.87) | 48.6 | 0.03 | 0.82 (0.64 to 1.04) | 0.09 |
| >1000 | 22 | 12237/19347 (63.2) | 12272/19261 (63.7) | 0.99 (0.92 to 1.06) | 16.0 | 0.25 | Referent |  |
| Trial duration, months |  |  |  |  |  |  |  |  |
| ≤12 | 27 | 1667/4357 (38.3) | 1711/4110 (41.6) | 0.81 (0.70 to 0.93) | 41.9 | 0.01 | 0.89 (0.72 to 1.10) | 0.28 |
| >12 | 7 | 11708/17401 (67.3) | 11699/17353 (67.4) | 0.99 (0.95 to 1.04) | 0.0 | 0.91 | Referent |  |

[a] Data from two trials that included higher-dose, lower-dose and placebo arms<sup>21,48</sup> are excluded, since the higher-dose and lower-dose arms in these studies spanned the 1,000 IU/day cut-off, rendering them unclassifiable for the purposes of this analysis. [b] Within-sub-group odds ratios from random effects model adjusting for study weights. [c] adjusted odds ratios and P values for interaction from multivariable meta-regression model including dichotomised variables for dose frequency, size and trial duration.

**Table S4:** Summary of Findings Table

| **Vitamin D<sub>3</sub> compared to placebo for prevention of acute respiratory infection (ARI)** |  |  |  |  |  |
| --- | --- | --- | --- | --- | --- |
| **Population:** children and adults of any age, sex or ethnic origin, with or without co-morbidity<br>**Setting:** Eighteen countries on four continents (Asia, Australasia, Europe, North America)<br>**Intervention:** oral vitamin D<sub>3</sub> (cholecalciferol) supplementation<br>**Comparison:** oral placebo |  |  |  |  |  |
| Outcomes | Anticipated absolute effects* (95% CI) |  | Relative effect (95% CI) | No of participants (studies) | Quality of the evidence (GRADE) |
|  | Risk with placebo | Risk with Vitamin D |  |  |  |
| Proportion with at least one ARI, all participants | 625 per 1,000 | 597 per 1,000 (574 to 620) | OR 0.91 (0.84 to 0.99) | 44009 (36 RCTs) | ⊕⊕⊕ MODERATE |
| Proportion with at least one ARI, participants in placebo-controlled trials with duration ≤12 months investigating daily dosing with 400-1,000 IU vitamin D<sub>3</sub>/day | 572 per 1,000 | 436 per 1,000 (375 to 500) | OR 0.58 (0.45 to 0.75) | 1232 (8 RCTs) | ⊕⊕⊕ MODERATE |
| Proportion with at least one hospital admission or emergency department attendance due to ARI | 13 per 1,000 | 12 per 1,000 (9 to 15) | OR 0.90 (0.70 to 1.16) | 19656 (18 RCTs) | ⊕⊕⊕ MODERATE |
| Proportion with serious adverse event, any cause | 42 per 1,000 | 40 per 1,000 (36 to 45) | OR 0.97 (0.86 to 1.09) | 27187 (35 RCTs) | ⊕⊕⊕ MODERATE |
| Proportion of deaths due to ARI or respiratory failure | 1 per 1,000 | 1 per 1,000 (1 to 1) | OR 1.04 (0.61 to 1.78) | 26670 (33 RCTs) | ⊕⊕⊕ MODERATE |
| *The risk in the intervention group (and its 95% confidence interval) is based on the assumed risk in the comparison group and the relative effect of the intervention (and its 95% CI).<br>CI: Confidence interval; OR: Odds ratio |  |  |  |  |  |
| **GRADE Working Group grades of evidence**<br>**High quality:** We are very confident that the true effect lies close to that of the estimate of the effect<br>**Moderate quality:** We are moderately confident in the effect estimate: The true effect is likely to be close to the estimate of the effect, but there is a possibility that it is substantially different<br>**Low quality:** Our confidence in the effect estimate is limited: The true effect may be substantially different from the estimate of the effect<br>**Very low quality:** We have very little confidence in the effect estimate: The true effect is likely to be substantially different from the estimate of effect |  |  |  |  |  |

**Table S5:**
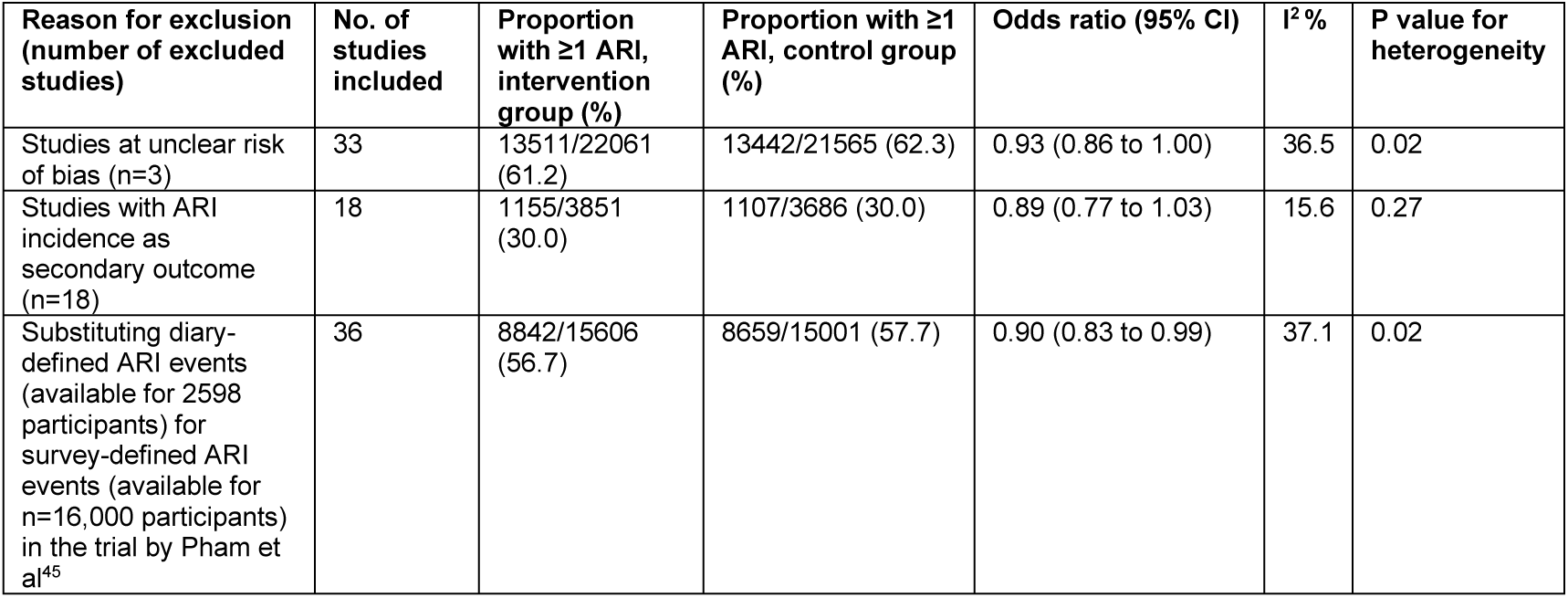
Results of exploratory sensitivity analyses excluding placebo-controlled trials at unclear risk of bias and studies investigating ARI incidence as a secondary outcome.

| Reason for exclusion (number of excluded studies) | No. of studies included | Proportion with $\geq 1$ ARI, intervention group (%) | Proportion with $\geq 1$ ARI, control group (%) | Odds ratio (95% CI) | I<sup>2</sup> % | P value for heterogeneity |
| --- | --- | --- | --- | --- | --- | --- |
| Studies at unclear risk of bias (n=3) | 33 | 13511/22061 (61.2) | 13442/21565 (62.3) | 0.93 (0.86 to 1.00) | 36.5 | 0.02 |
| Studies with ARI incidence as secondary outcome (n=18) | 18 | 1155/3851 (30.0) | 1107/3686 (30.0) | 0.89 (0.77 to 1.03) | 15.6 | 0.27 |
| Substituting diary-defined ARI events (available for 2598 participants) for survey-defined ARI events (available for n=16,000 participants) in the trial by Pham et al<sup>45</sup> | 36 | 8842/15606 (56.7) | 8659/15001 (57.7) | 0.90 (0.83 to 0.99) | 37.1 | 0.02 |

**Figure S1:**
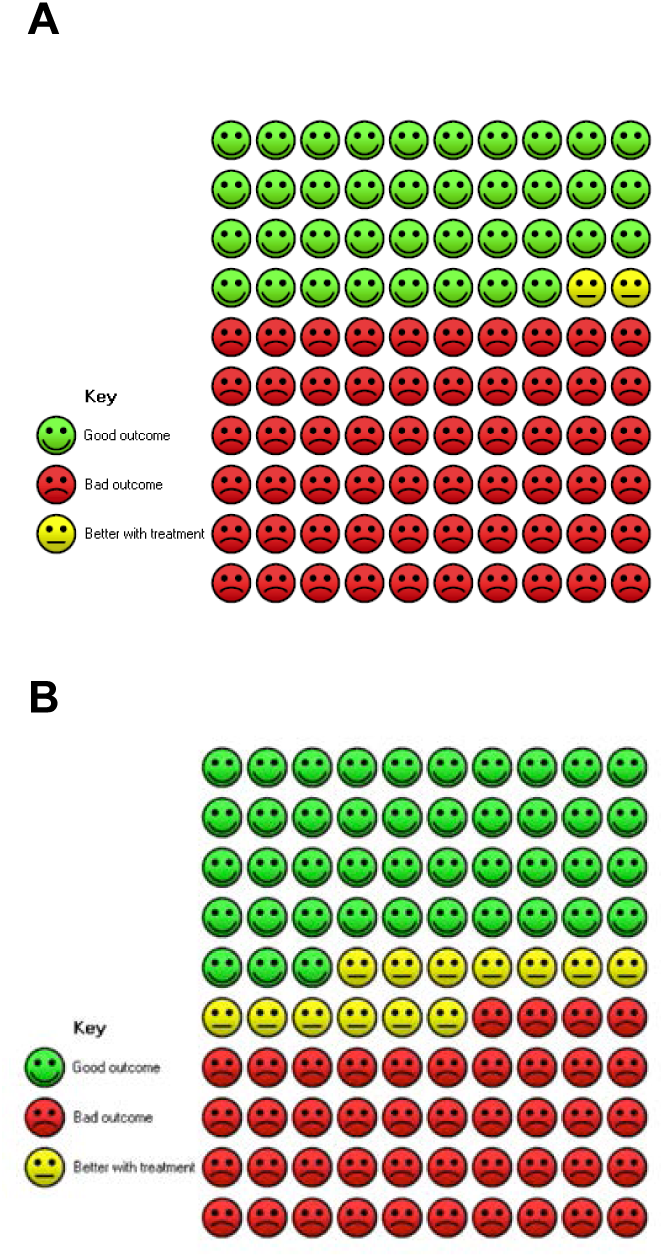
Cates plot illustrating reduction in risk of one or more acute respiratory infections with vitamin D supplementation vs. placebo (primary comparison), **A)** Overall, and **B)** in trials with duration ≤12 months where vitamin D_3_ was administered using daily doses of 400-1000 IU/day.

**Figure S2:**
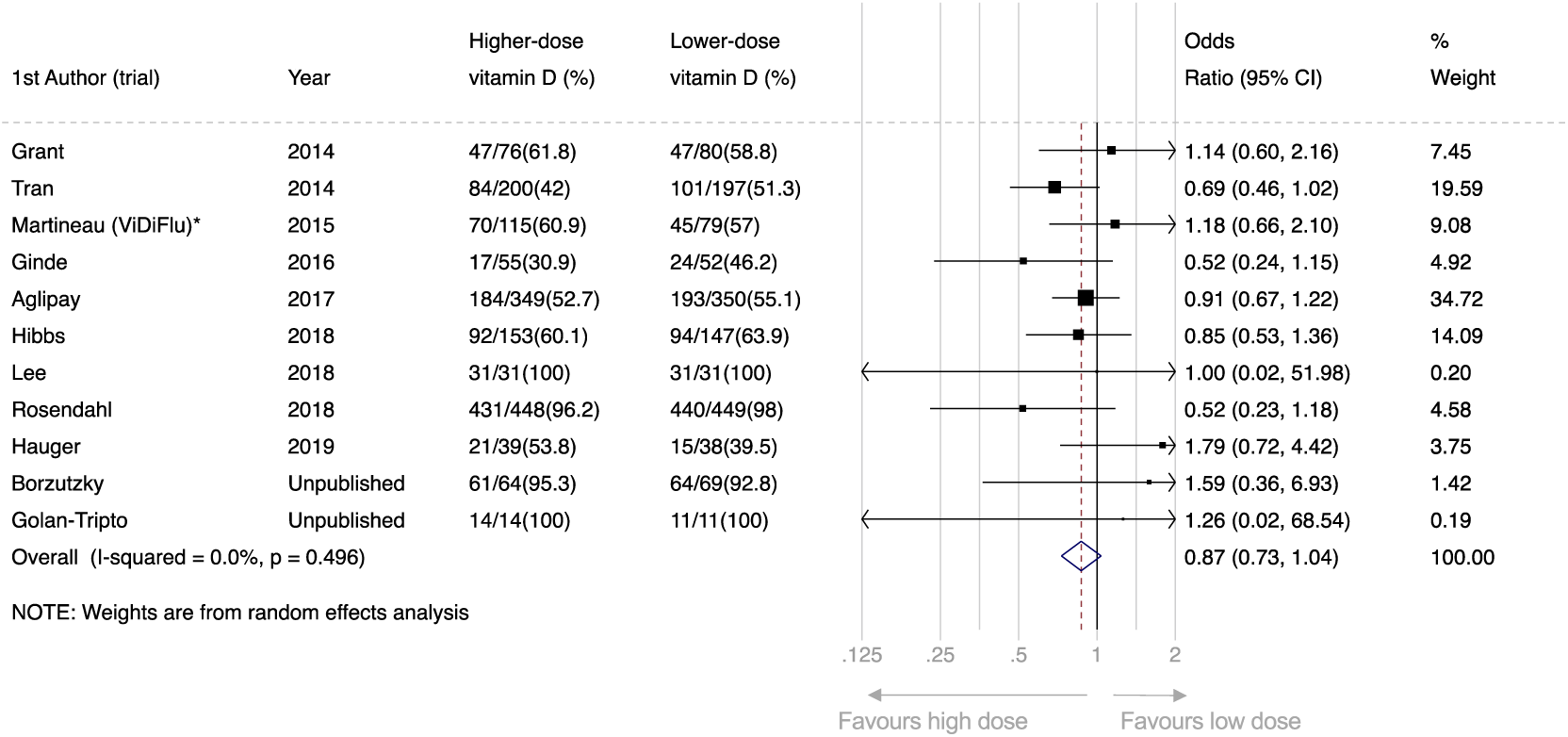
Forest plot of RCTs comparing effects of higher-vs. lower-dose vitamin D, reporting proportion of participants experiencing at least one acute respiratory infection. *This analysis includes data from the subset of ViDiFlu trial participants who were randomised to higher-vs. lower-dose vitamin D.

**Figure S3:**
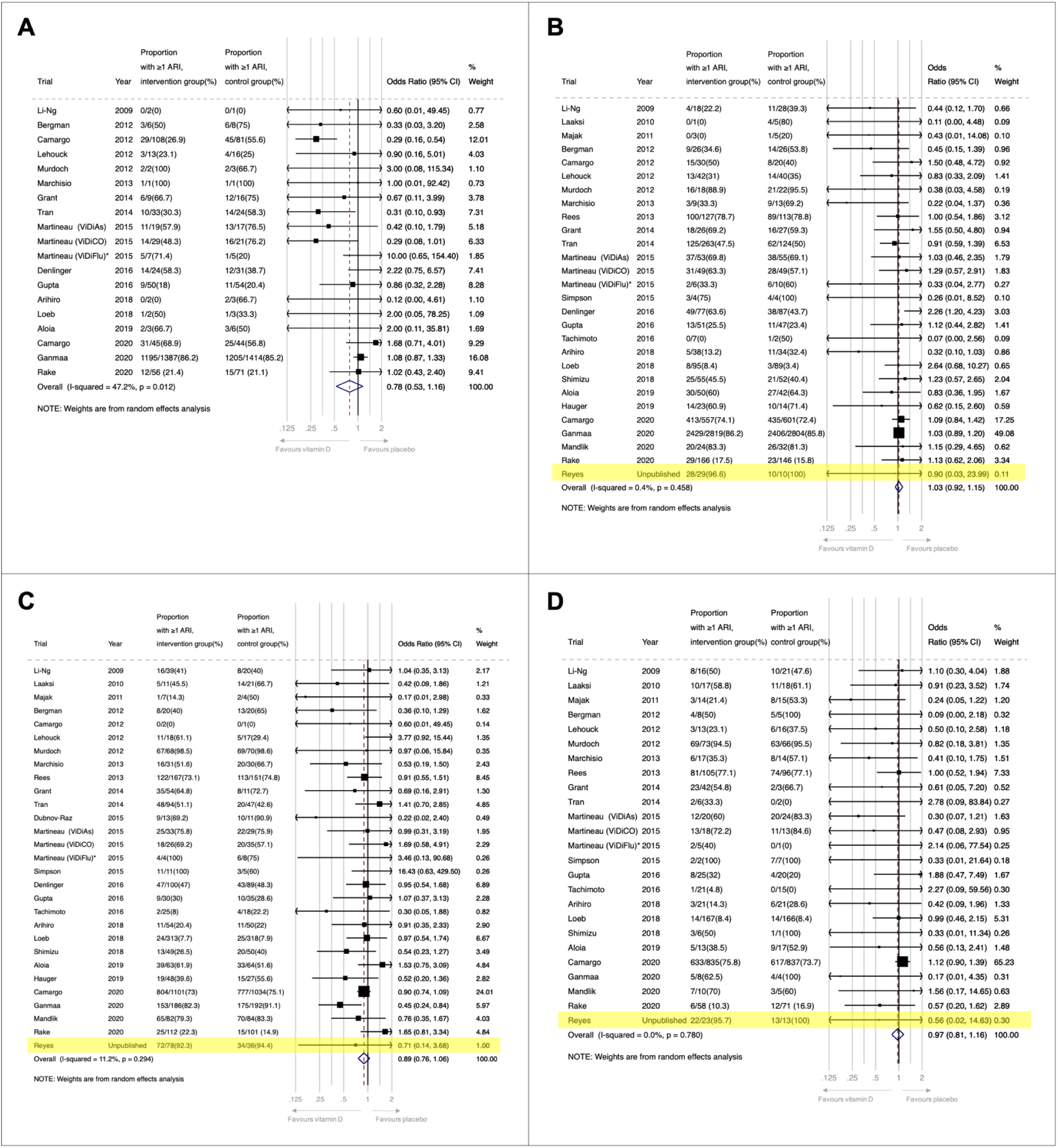
Forest plot of RCTs comparing effects of vitamin D vs. placebo, reporting proportion of participants experiencing at least one acute respiratory infection, by baseline 25-hydroxyvitamin D level. **A)** <25.0 nmol/L; **B)** 25.0 to 49.9 nmol/L; **C)** 50.0 to 74.9 nmol/L, and **D)** ≥75.0 nmol/L.

**Figure S4:**
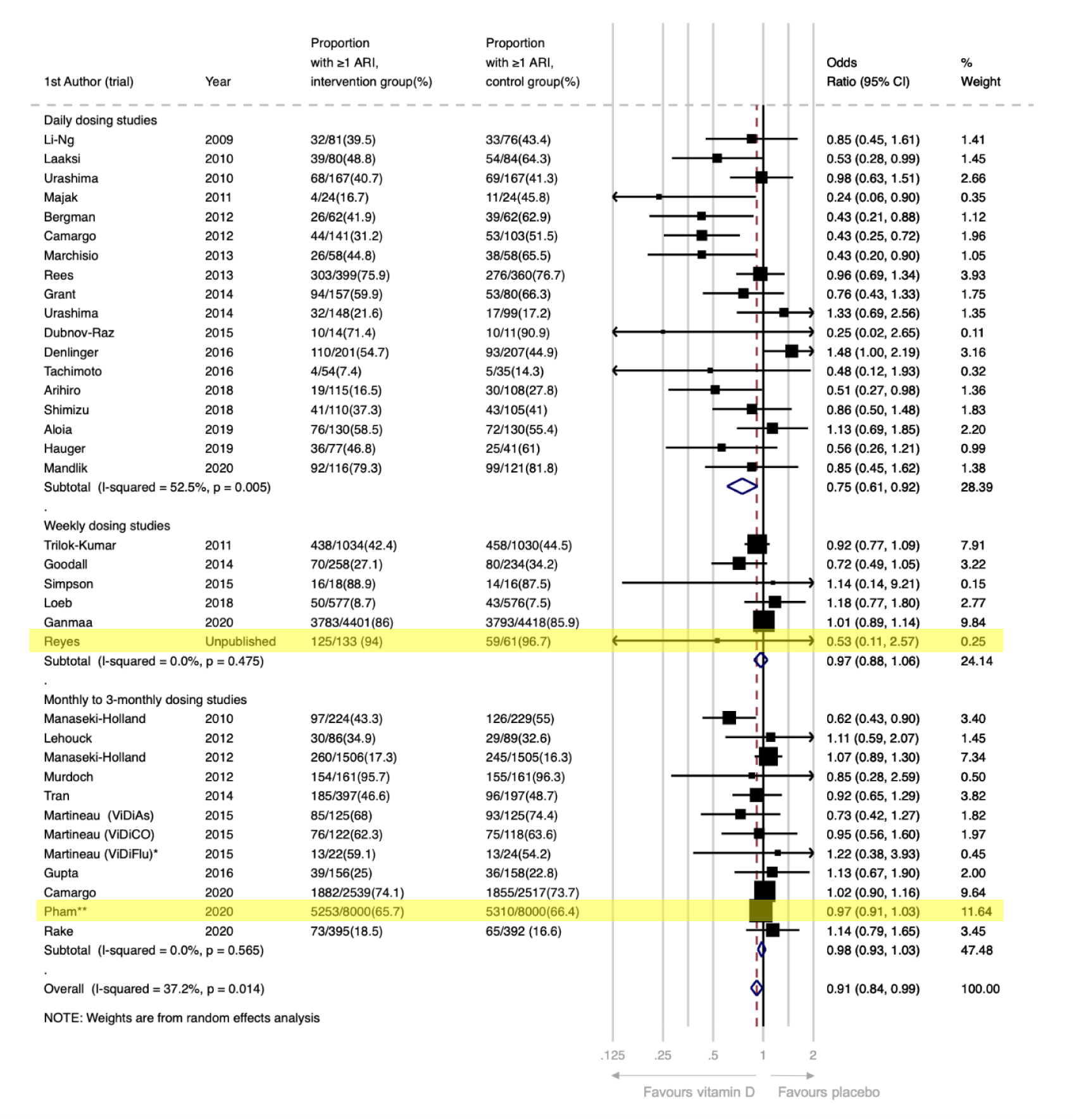
Forest plot of RCTs comparing effects of vitamin D vs. placebo, reporting proportion of participants experiencing at least one acute respiratory infection, by frequency of supplementation (daily vs. weekly vs. monthly to 3-monthly) *This analysis includes data from the subset of ViDiFlu trial participants who were randomised to vitamin D vs. placebo control. **For this trial, participants were asked to report the occurrence of ARTI during the one month prior to completing each annual survey (max surveys=5). The numerator is the number of people who reported an ARTI on at least one survey. The ARTI outcomes for people who completed fewer than 5 surveys and who did not report an ARTI (N=2239; 14%) were estimated based on the % affected among those who completed all 5 surveys (N=12,152; 76%).

**Figure S5:**
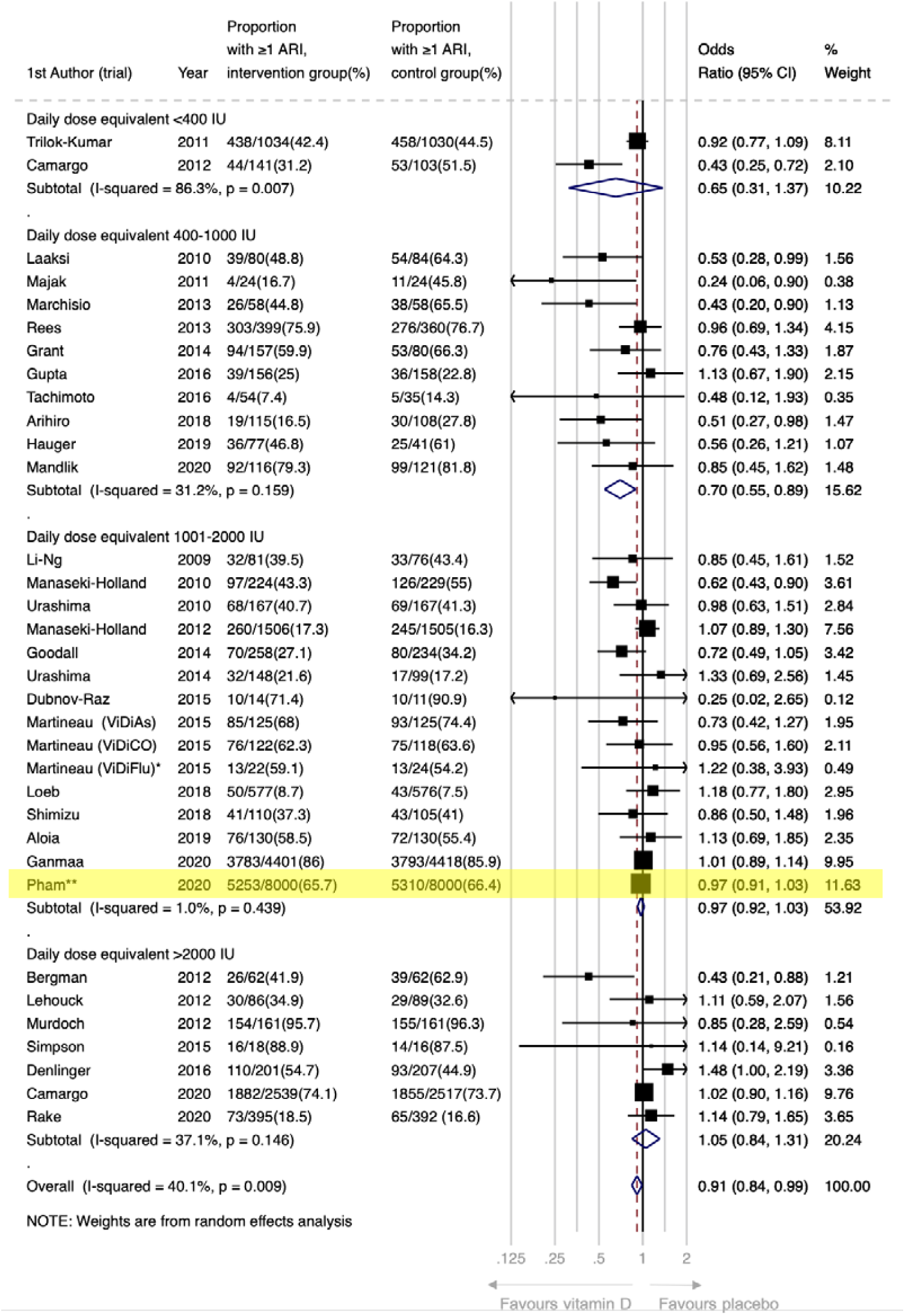
Forest plot of RCTs comparing effects of vitamin D vs. placebo, reporting proportion of participants experiencing at least one acute respiratory infection, by daily dose equivalents (<400 IU/day vs. 400-1000 IU/day vs. 1001-2000 IU/day vs. >2000 IU/day). *This analysis includes data from the subset of ViDiFlu trial participants who were randomised to vitamin D vs. placebo control. **For this trial, participants were asked to report the occurrence of ARTI during the one month prior to completing each annual survey (max surveys=5). The numerator is the number of people who reported an ARTI on at least one survey. The ARTI outcomes for people who completed fewer than 5 surveys and who did not report an ARTI (N=2239; 14%) were estimated based on the % affected among those who completed all 5 surveys (N=12,152; 76%).

**Figure S6:**
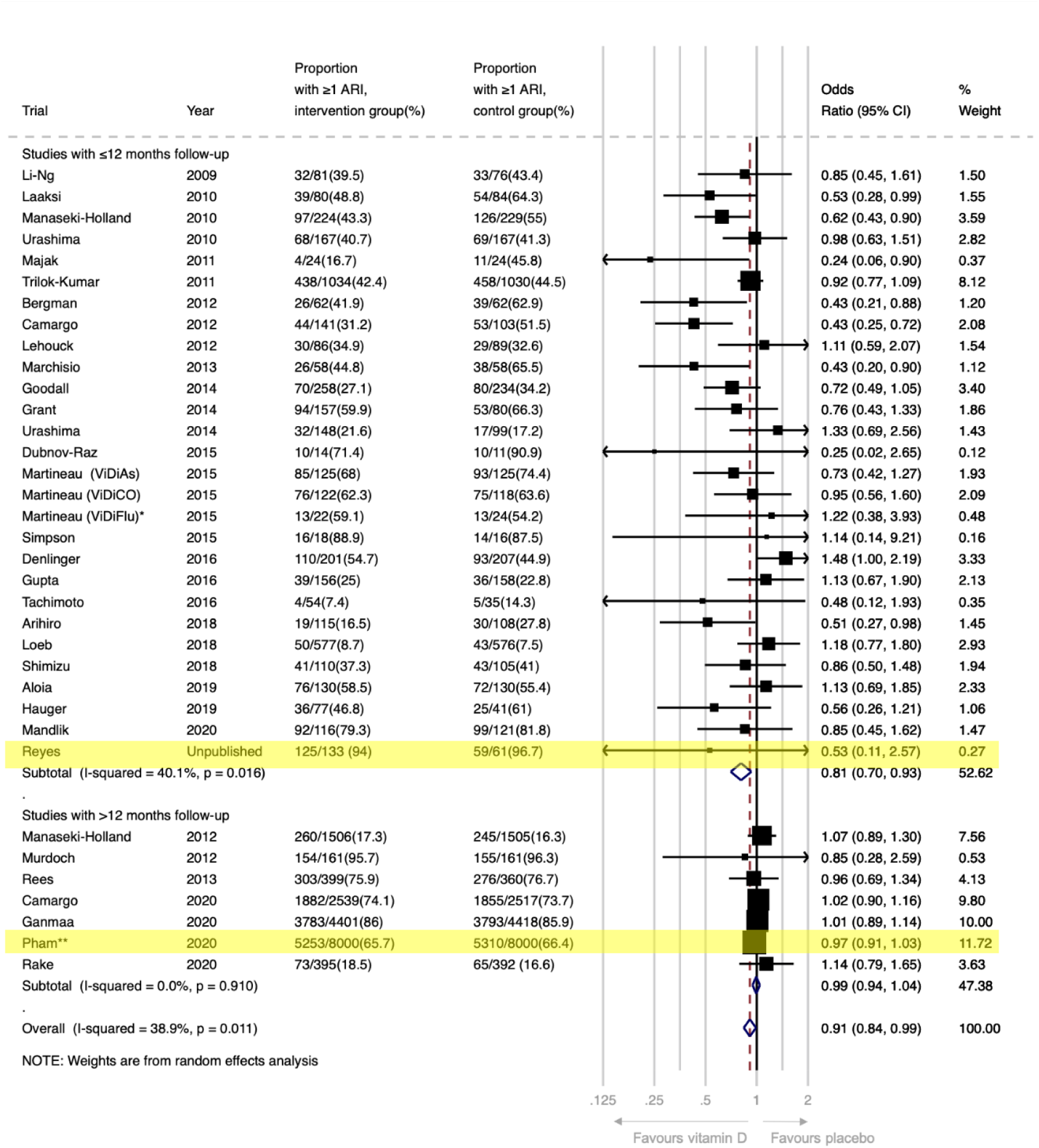
Forest plot of RCTs comparing effects of vitamin D vs. placebo, reporting proportion of participants experiencing at least one acute respiratory infection, by trial duration (≤12 months vs. >12 months). *This analysis includes data from the subset of ViDiFlu trial participants who were randomised to vitamin D vs. placebo control. **For this trial, participants were asked to report the occurrence of ARTI during the one month prior to completing each annual survey (max surveys=5). The numerator is the number of people who reported an ARTI on at least one survey. The ARTI outcomes for people who completed fewer than 5 surveys and who did not report an ARTI (N=2239; 14%) were estimated based on the % affected among those who completed all 5 surveys (N=12,152; 76%).

**Figure S7:**
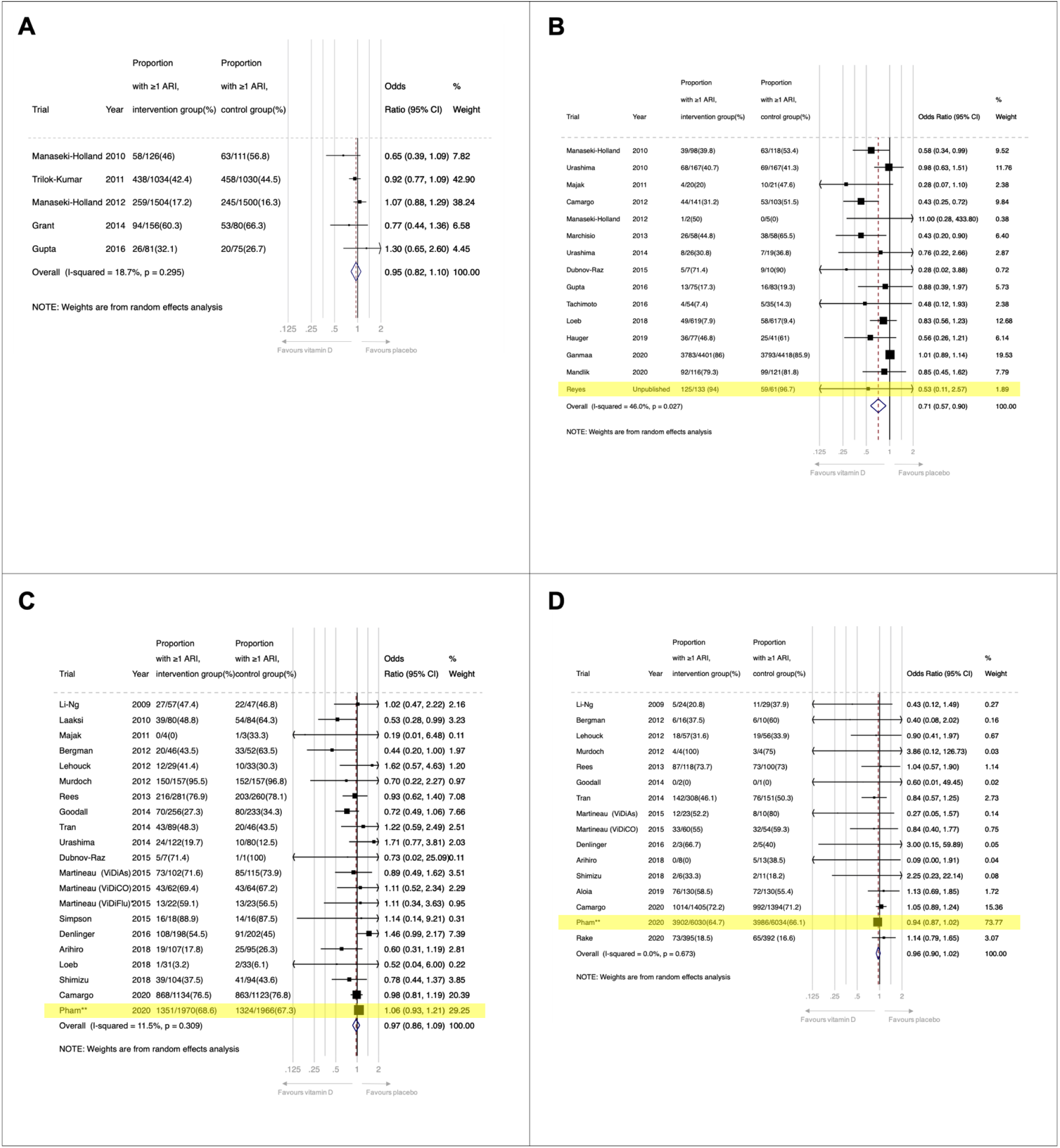
Forest plot of RCTs comparing effects of vitamin D vs. placebo, reporting proportion of participants experiencing at least one acute respiratory infection, by the following age strata: **A)** <1 year; **B)** 1-15.99 years; **C)** 16-64.99 years, and **D)** ≥65 years *This analysis includes data from the subset of ViDiFlu trial participants who were randomised to vitamin D vs. placebo control. **For this trial, participants were asked to report the occurrence of ARTI during the one month prior to completing each annual survey (max surveys=5). The numerator is the number of people who reported an ARTI on at least one survey. The ARTI outcomes for people who completed fewer than 5 surveys and who did not report an ARTI (N=2239; 14%) were estimated based on the % affected among those who completed all 5 surveys (N=12,152; 76%).

**Figure S8:**
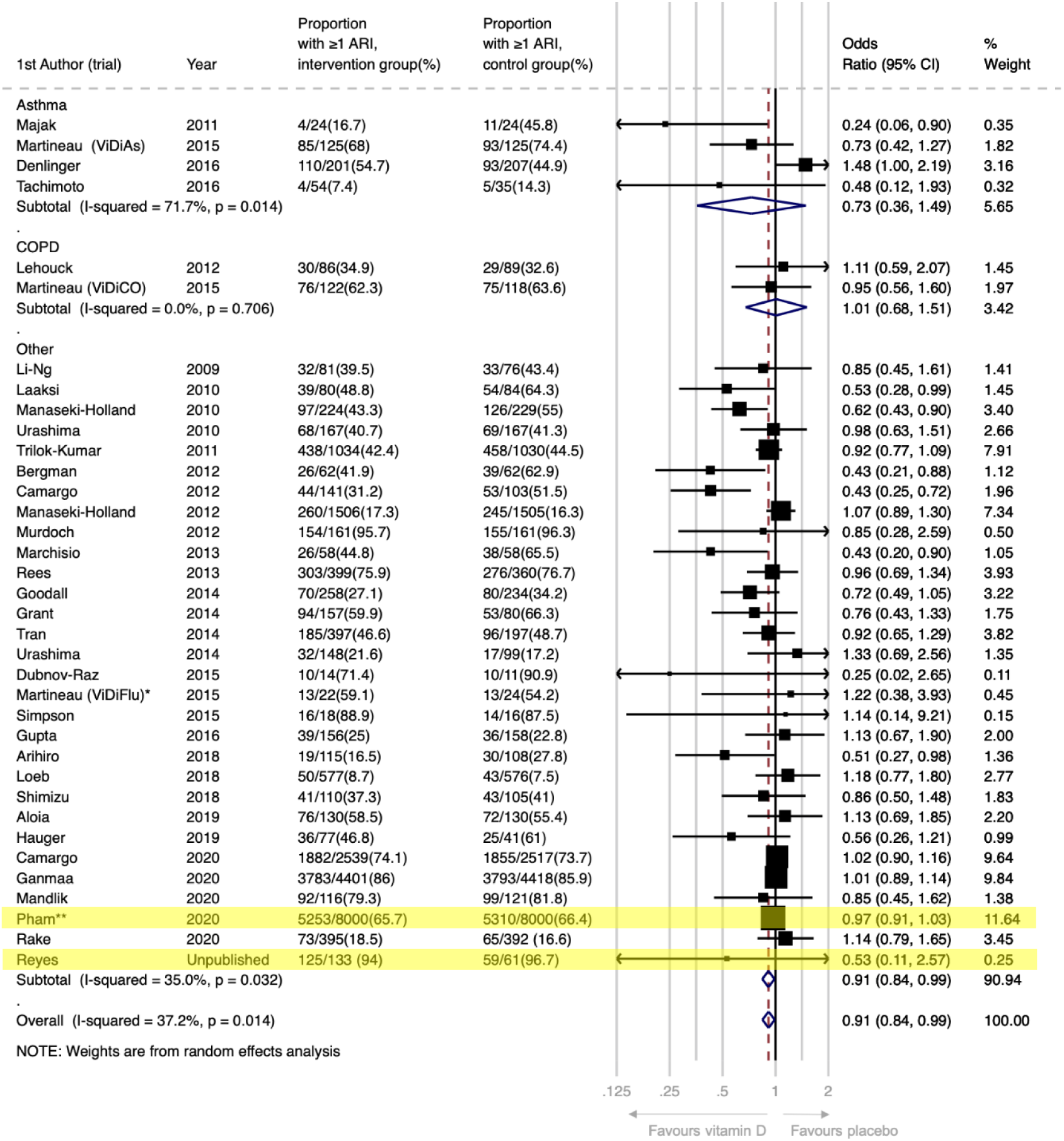
Forest plot of RCTs comparing effects of vitamin D vs. placebo, reporting proportion of participants experiencing at least one acute respiratory infection, by presence or absence of airway disease comorbidity. *This analysis includes data from the subset of ViDiFlu trial participants who were randomised to vitamin D vs. placebo control. **For this trial, participants were asked to report the occurrence of ARTI during the one month prior to completing each annual survey (max surveys=5). The numerator is the number of people who reported an ARTI on at least one survey. The ARTI outcomes for people who completed fewer than 5 surveys and who did not report an ARTI (N=2239; 14%) were estimated based on the % affected among those who completed all 5 surveys (N=12,152; 76%).

**Figure S9.**
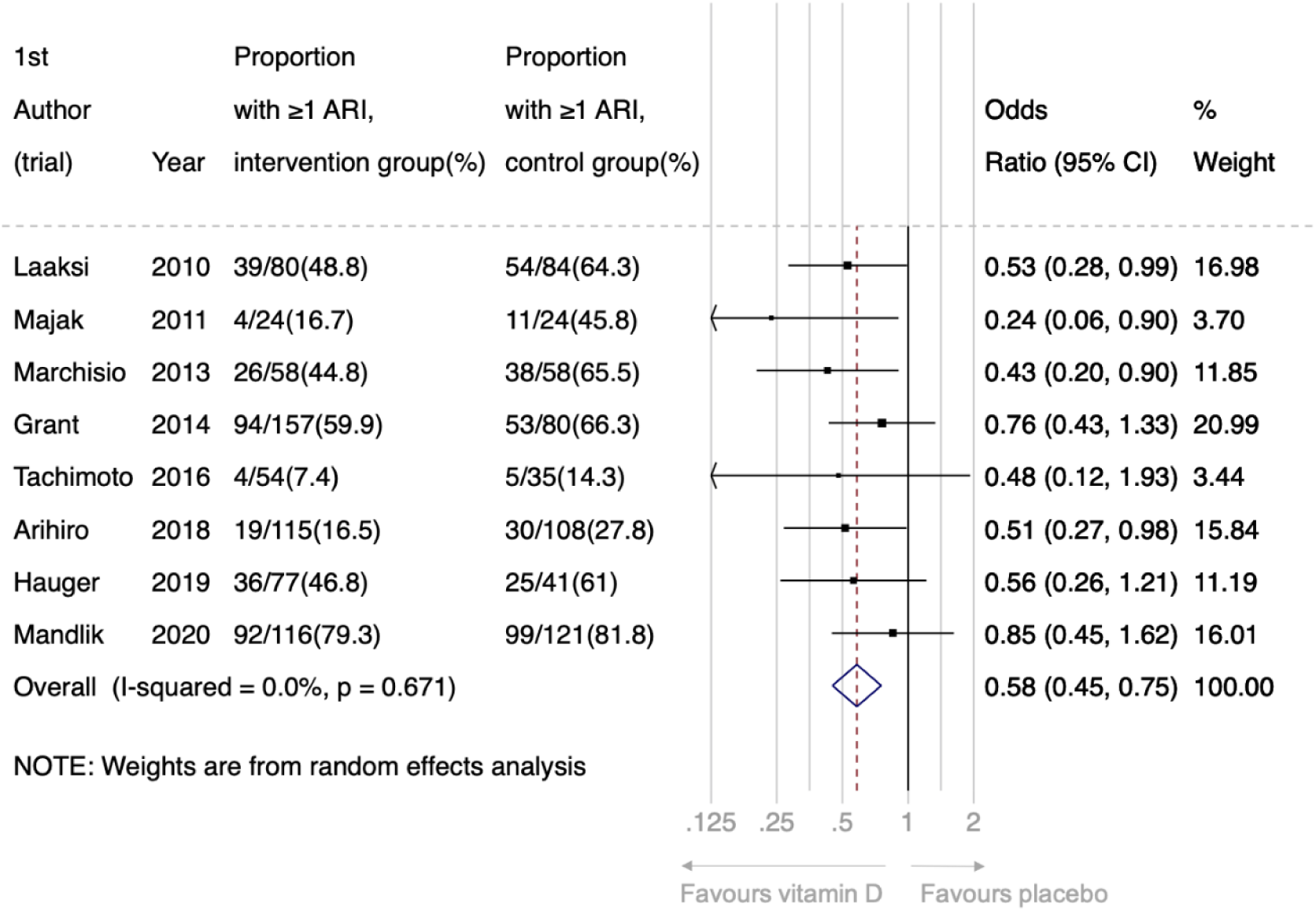
Forest plot of sub-set of RCTs with duration ≤12 months comparing effects of daily vitamin D at a dose of 400-1000 IU/day vs. placebo, reporting proportion of participants experiencing at least one acute respiratory infection

**Figure S10:**
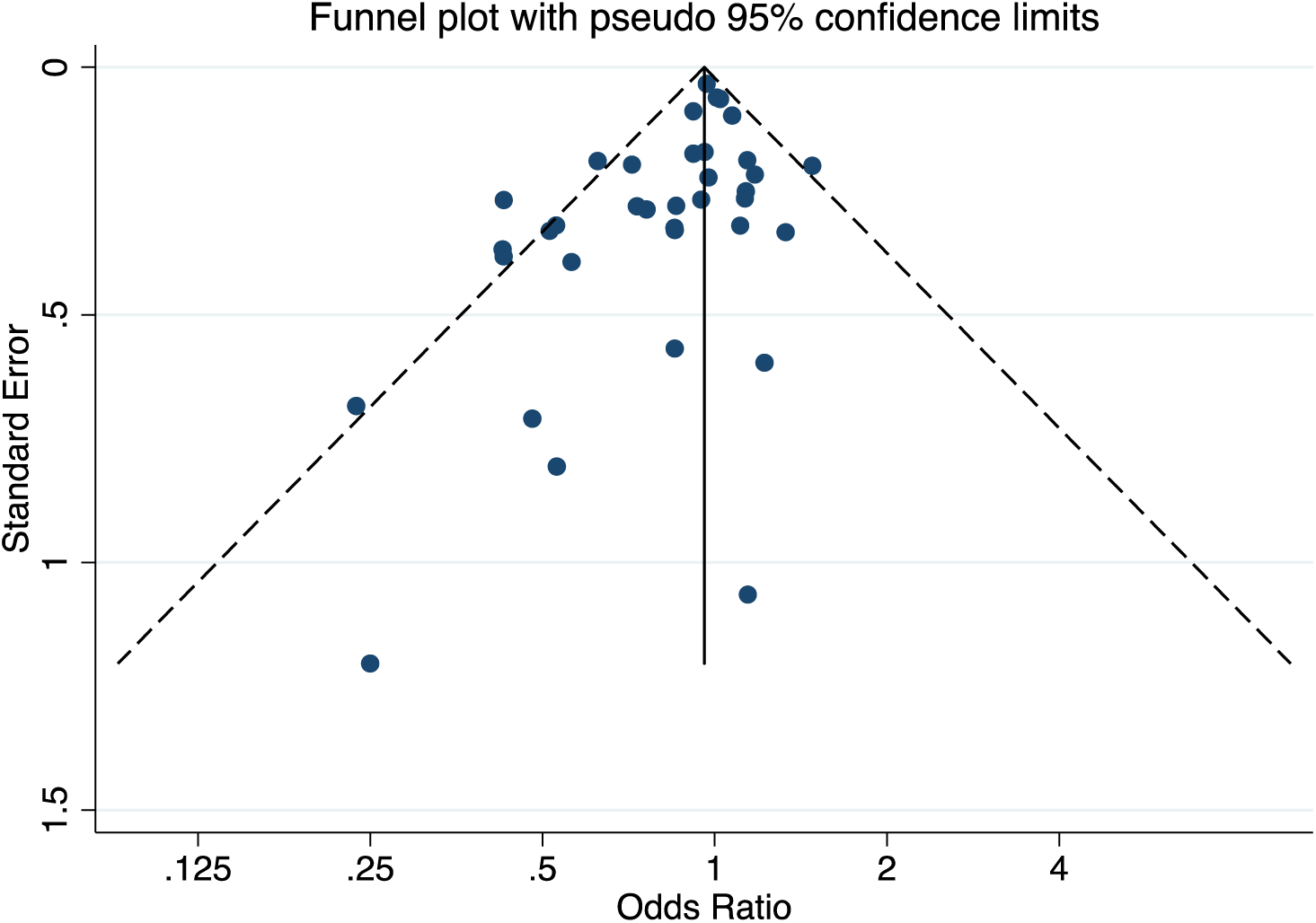
Funnel plot of placebo-controlled RCTs reporting proportion of participants experiencing 1 or more acute respiratory infection. Egger’s test for publication bias: P=0.008

